# Characteristics and outcomes of 118,155 COVID-19 individuals with a history of cancer in the United States and Spain

**DOI:** 10.1101/2021.01.12.21249672

**Authors:** Elena Roel, Andrea Pistillo, Martina Recalde, Anthony G. Sena, Sergio Fernández-Bertolín, María Aragón, Diana Puente, Waheed-Ul-Rahman Ahmed, Heba Alghoul, Osaid Alser, Thamir M. Alshammari, Carlos Areia, Clair Blacketer, William Carter, Paula Casajust, Aedin C. Culhane, Dalia Dawoud, Frank DeFalco, Scott L. Duvall, Thomas Falconer, Asieh Golozar, Mengchun Gong, Laura Hester, George Hripcsak, Eng Hooi Tan, Hokyun Jeon, Jitendra Jonnagaddala, Lana Y.H. Lai, Kristine E. Lynch, Michael E. Matheny, Daniel R. Morales, Karthik Natarajan, Fredrik Nyberg, Anna Ostropolets, José D. Posada, Albert Prats-Uribe, Christian G. Reich, Donna Rivera, Lisa M. Schilling, Isabelle Soerjomataram, Karishma Shah, Nigam Shah, Yang Shen, Matthew Spotniz, Vignesh Subbian, Marc A. Suchard, Annalisa Trama, Lin Zhang, Ying Zhang, Patrick Ryan, Daniel Prieto-Alhambra, Kristin Kostka, Talita Duarte-Salles

**Affiliations:** Fundació Institut Universitari per a la recerca a l’Atenció Primària de Salut Jordi Gol i Gurina (IDIAPJGol), Barcelona, Spain; Universitat Autònoma de Barcelona, Spain; Janssen Research and Development, Titusville, NJ USA; Department of Medical Informatics, Erasmus University Medical Center, Rotterdam, The Netherlands; NDORMS, University of Oxford, Botnar Research Centre, Windmill Road, Oxford, OX3 7LD, UK; College of Medicine and Health, University of Exeter, St Luke’s Campus, Heavitree Road, Exeter, EX1 2LU, UK; Faculty of Medicine, Islamic University of Gaza, Palestine; Massachusetts General Hospital, Harvard Medical School, USA; Medication Safety Research Chair, King Saud University; Nuffield Department of Clinical Neurosciences, University of Oxford, Oxford, OX3 9DU, UK; Data Science to Patient Value Program, Department of Medicine, University of Colorado Anschutz Medical Campus; Real-World Evidence, Trial Form Support, Barcelona, Spain; Department of Data Science, Dana-Farber Cancer Institute, Boston MA, USA; Department of Biostatistics, Harvard TH Chan School of Public Health, Boston MA, USA; Faculty of Pharmacy, Cairo University, Cairo, Egypt; VA Informatics and Computing Infrastructure, VA Salt Lake City Health Care System, Salt Lake City, UT, USA; Department of Internal Medicine, University of Utah School of Medicine, Salt Lake City, UT, USA; Department of Biomedical Informatics, Columbia University, New York, NY, US; New York-Presbyterian Hospital, 622 W 168 St, PH20 New York, NY 10032 USA; Department of Epidemiology, Johns Hopkins School of Public, Baltimore MD, USA; Pharmacoepidemiology, Regeneron Pharmaceuticals, NY, US; Digital Health China Technologies Co., LTD, Beijing, China; Associate Director, Epidemiology, Janssen Research and Development, LLC; Centre for Statistics in Medicine, NDORMS, University of Oxford, OX3 7LD, UK; Department of Biomedical Sciences, Ajou University Graduate School of Medicine, Suwon, Gyeonggi-do, Republic of Korea; School of Public Health and Community Medicine, UNSW Sydney; School of Medical Sciences, University of Manchester, UK; Tennessee Valley Healthcare System, Veterans Affairs Medical Center, Nashville, TN, USA; Department of Biomedical Informatics, Vanderbilt University Medical Center, Nashville, TN, USA; Division of Population Health and Genomics, University of Dundee, UK; University of Southern Denmark; School of Public Health and Community Medicine, Institute of Medicine, Sahlgrenska Academy, University of Gothenburg, Gothenburg, Sweden; Department of Medicine, School of Medicine, Stanford University, Redwood City, CA USA; Real World Solutions, IQVIA, Cambridge, MA USA; Division of Cancer Control and Population Sciences, National Cancer Institute, Rockville, MD; Section of Cancer Surveillance, International Agency for Research on Cancer, 150 Cours Albert Thomas, 69008 Lyon, France; College of Engineering, The University of Arizona, Tucson, Arizona, USA; Fielding School of Public Health, University of California, Los Angeles; Fondazione IRCSS Istituto Nazionale dei Tumori, Milan – Italy; School of Population Medicine and Public Health, Peking Union Medical College, Chinese Academy of Medical Sciences; School of Population Health and Global Health, The University of Melbourne

**Keywords:** COVID-19, SARS-CoV-2, cancer, characterization, hospital admission, mortality

## Abstract

**Purpose:** We aimed to describe the demographics, cancer subtypes, comorbidities and outcomes of patients with a history of cancer with COVID-19 from March to June 2020. Secondly, we compared patients *hospitalized* with COVID-19 to patients *diagnosed* with COVID-19 and patients *hospitalized* with influenza.

**Methods:** We conducted a cohort study using eight routinely-collected healthcare databases from Spain and the US, standardized to the Observational Medical Outcome Partnership common data model. Three cohorts of patients with a history of cancer were included: i) *diagnosed* with COVID-19, ii) *hospitalized* with COVID-19, and iii) *hospitalized* with influenza in 2017-2018. Patients were followed from index date to 30 days or death. We reported demographics, cancer subtypes, comorbidities, and 30-day outcomes.

**Results:** We included 118,155 patients with a cancer history in the COVID-19 *diagnosed* and 41,939 in the COVID-19 *hospitalized* cohorts. The most frequent cancer subtypes were prostate and breast cancer (range: 5-19% and 1-14% in the *diagnosed* cohort, respectively). Hematological malignancies were also frequent, with non-Hodgkin’s lymphoma being among the 5 most common cancer subtypes in the *diagnosed* cohort. Overall, patients were more frequently aged above 65 years and had multiple comorbidities. Occurrence of death ranged from 8% to 14% and from 18% to 26% in the *diagnosed* and h*ospitalized* COVID-19 cohorts, respectively. Patients hospitalized with influenza (n=242,960) had a similar distribution of cancer subtypes, sex, age and comorbidities but lower occurrence of adverse events.

**Conclusion:** Patients with a history of cancer and COVID-19 have advanced age, multiple comorbidities, and a high occurence of COVID-19-related events. Additionaly, hematological malignancies were frequent in these patients.This observational study provides epidemiologic characteristics that can inform clinical care and future etiological studies.

## Background

Shortly after the emergence of the novel coronavirus disease 2019 (COVID-19), patients with cancer were reported to be a high-risk population for COVID-19.^1,2^ These patients have an increased susceptibility to infections as a result of their immunosuppressed state, caused by the cancer itself, certain types of chemo- or immunotherapy, or surgery and a higher exposure to healthcare-associated infections (HAI).^3^ In addition, patients with cancer are often older and with additional comorbidities, which might increase their risk of worse COVID-19 outcomes.^4^

Prior studies assessing COVID-19-related risks in the cancer population have demonstrated conflicting results. Some studies found that patients with cancer have an increased risk of COVID-19 related hospitalization, admission to intensive care units, and mortality compared to patients without cancer, ^1,2,4,5^ whereas others did not.^6,7^ These studies included a limited number of patients with cancer, were conducted across different countries and geographic regions with different incidence of infection at varying time points and used different definitions for cancer (e.g., active cancer, solid cancer, history of cancer), which limit their generalizability. Furthermore, they presented results for models adjusted by (different) arbitrary covariates, without a theoretical framework of confounding variables, which limits the interpretation for descriptive and causal inference purposes.^8,9^

Given that COVID-19 is a novel disease, large descriptive studies are essential to inform public health strategies and clinical care, as well as to provide the groundwork for future etiological studies. Large studies with detailed information of medical conditions and health outcomes, such as thromboembolic events, in patients with cancer and COVID-19 are lacking to date. To fill that gap, we conducted a multi-national study aiming to describe the demographics, cancer subtypes, comorbidities and outcomes of patients with a history of cancer and COVID-19. In addition, we compared patients with a history of cancer hospitalized with COVID-19 to i) patients with a history of cancer diagnosed with COVID-19; and ii) patients with a history of cancer hospitalized with seasonal influenza (2017-2018) as a benchmark.

## Methods

### Study design, setting and data sources

This study was part of the CHARYBDIS (Characterizing Health Associated Risks, and Your Baseline Disease In SARS-COV-2) project designed by the Observational Health Data Sciences and Informatics (OHDSI) community. CHARYBDIS is large-scale study aiming to characterize individuals with COVID-19 using routinely-collected healthcare data (protocol available at https://www.ohdsi.org/wp-content/uploads/2020/07/Protocol_COVID-19-Charybdis-Characterisation_V5.docx). Twenty-two databases standardized to the Observational Medical Outcomes Partnership (OMOP) Common Data Model (CDM)^10^ have contributed to CHARYBDIS to date. The OHDSI network maintains the OMOP-CDM, and its members have developed a wide range of tools to facilitate analyses of such mapped data.^11^ Results for this sub-study were extracted from the overarching result set on October, 9th, 2020.

From the databases that contributed to CHARYBDIS, we included those reporting data on at least 140 subjects with a history of cancer diagnosed and/or hospitalized with COVID-19. This cut-off was established to estimate the prevalence of conditions and 30-days outcomes affecting 10% of the study population with a confidence interval (CI) width of +/-5%. The selection process is depicted in Appendix Figure A1. Up to eight databases from two countries (Spain and the United States (US)) were included in this study. Spanish data came from SIDIAP, a primary care database linked to hospital admissions.^12^ Data from the US included Electronic Health Records (EHR) from the hospital setting: CU-AMC-HDC, CUIMC, Optum-EHR,^13^ and VA-OMOP (93% male, mostly veterans); as well as claims data: HealthVerity and IQVIA-OpenClaims. All databases reported patients with COVID-19 identified from March to June 2020, apart from CU-AMC-HDC that included data to August 2020. For a description of the included data sources, see Appendix Table A1.

### Study participants

We included three non-mutually exclusive cohorts of patients with a history of cancer: i) patients *diagnosed* with COVID-19, ii) patients *hospitalized* with COVID-19, and iii) patients hospitalized with seasonal influenza in 2017-2018.

To comprehensively capture baseline characteristics, we only included patients with at least one year of observation time available prior to index date (i.e., date of start of the cohort). Patients with a history of cancer were defined as those having a record of any malignant neoplasm excluding non-melanoma skin cancer prior to the index date. Patients *diagnosed* with COVID-19 were those having a clinical diagnosis and/or a positive test for the severe acute respiratory syndrome coronavirus 2 (SARS-CoV-2) documented in outpatient or inpatient records. Patients *hospitalized* with COVID-19 were those who had a hospitalization episode and a COVID-19 clinical diagnosis or positive SARS-CoV-2 test within a time window of 21 days prior to admission up to the end of their hospitalization. We chose this time window to include patients with a diagnosis prior to hospitalization and to allow for a record delay in test results or diagnoses. Similarly, patients hospitalized with seasonal influenza were those who had a hospitalization episode and a influenza clinical diagnosis or positive test result for influenza in 2017-2018.^14^ The criteria to define patients with a history of cancer and COVID-19 and influenza cases can be found in Appendix Table A2.

Index date for the *diagnosed* cohort was the date of clinical diagnosis or the earliest test day registered within seven days of a first positive test, whichever occurred first. Index date for both *hospitalized* cohorts (COVID-19 and influenza) was the day of hospitalization. Patients were followed from the index date to the earliest of either death, end of the observation period,^15^ or 30 days.

### Patient characteristics and outcomes

More than 15,000 medical conditions from up to one year prior to index date were identified in each cohort based on the Systematized Nomenclature of Medicine (SNOMED) hierarchy, with all descendant codes included.^15^ In addition, we created specific definitions for comorbidities and outcomes of particular interest (available in Appendix Table A2). To describe the frequency of cancer subtypes by topographical location (henceforth, referred to as cancer types), we selected 26 cancer types based on the most prevalent cancers in Spain and the US, according to data from the International Agency for Research on Cancer.^16^ The SNOMED codes used to identify each cancer type can be found in Appendix Table A3. We report here demographics (sex and age), antineoplastic and immunomodulating treatment received in the month and year prior to the index date and a selection of key comorbidities (based on their relevance to the COVID-19 field and on their prevalence in the cohorts).

The 30-day outcomes of interest in the COVID-19 *diagnosed* cohort were hospitalization and death (from all causes). In the *hospitalized c*ohorts (COVID-19 and influenza), the outcomes of interest were acute respiratory distress syndrome (ARDS), acute kidney injury (AKI), cardiovascular disease events, deep vein thrombosis, pulmonary embolism, sepsis, requirement of intensive services (identified by a recorded mechanical ventilation and/or a tracheostomy and/or extracorporeal membrane oxygenation procedure) and death (from all causes). SIDIAP only reported death and hospitalization, whereas CU-AMC-HDC did not report any outcome.

### Data characterization and analysis

Analysis was performed through a federated analysis approach.^15^ Following a pre-specified analysis plan, a common analytical code for the whole CHARYBDIS study was developed by the OHDSI Methods library and run locally in each participating site (code available at zenodo.org).^17^ Individual-level data remained within host institutions and only aggregate results were provided to the research team and publicly-shared. The results reported in this manuscript, and additional data, are available for consultation on a regularly updated website as new databases and/or results are added (https://data.ohdsi.org/Covid19CharacterizationCharybdis/).

Herein, we describe the lifetime cancer prevalence (recorded any time prior to the index date) and the number of patients included by database and cohort. Results are calculated and reported by cohort and database. Demographics, cancer types, comorbidities, and outcomes are reported as proportions (calculated as the number of persons within a given category, divided by the total number of persons), along with 95%CI. To calculate these proportions, a minimum count required of 5 individuals was established to minimize the risk of re identification of patients.We also report the ranking of the 10 most common cancer types by frequency. In addition, we summarized the prevalence of all the baseline conditions retrieved in a Manhattan-style plot.

To compare characteristics between study cohorts, we calculated standardized mean differences (SMD), with an SMD >|0.1| indicating a meaningful difference in the prevalence of a given condition.^18^ As this study was designed as a detailed descriptive study, statistical modelling was out of scope in the developed analytical packages. Therefore, differences across the groups compared should not be interpreted as causal effects.

We used R version 3.6 for data visualization. All the data partners obtained Institutional Review Board (IRB) approval or exemption to conduct this study.

## Results

### Lifetime cancer prevalence

Overall, the databases included in this study identified 906,691 patients *diagnosed* with COVID-19 and 196,018 patients *hospitalized* with COVID-19. The lifetime cancer prevalence ranged from 4% (HealthVerity) to 25% (STARR-OMOP) in all patients *diagnosed* with COVID-19; whereas it ranged from 11% (HealthVerity) to 40%(VA-OMOP) in all patients *hospitalized* with COVID-19 (Appendix Table A4). In addition, 242,960 patients hospitalized with seasonal influenza in 2017-2018 were identified. Among those, the lifetime cancer prevalence ranged from 21% (Optum-EHR) to 39% (VA-OMOP).

We included 118,155 patients *diagnosed* (Spain: 8,854; US: 109,301) and 41,939 patients *hospitalized* (Spain: 2,610; US: 39,329) with COVID-19 and history of cancer and 62,077 patients hospitalized (all from the US) with seasonal influenza and history of cancer.

### Demographics

The distribution of demographics, comorbidities and outcomes of both COVID-19 cohorts can be found in Table 1 (95%CI of each condition available in Appendix Table A5). In the *diagnosed* cohort, patients were more commonly female (range 53% to 55%), aside from STARR-OMOP (47%) and VA-OMOP (7.1%). In contrast, in the *hospitalized cohort*, male predominated in all databases (range 50% to 60%, VA-OMOP: 96%). Overall, patients were mainly aged above 65 years in both COVID-19 *diagnosed* and *hospitalized* cohorts and patients *hospitalized* were consistently older than those *diagnosed* (Appendix Figure A2). In the *diagnosed* cohort, the proportion of patients having received antineoplastic and immunomodulating agents the month and the year prior to index date ranged from 5% (HealthVerity) to 23% (STARR-OMOP) and from 14% (SIDIAP) to 35% (CU-AMC-DHC), respectively. Similarly, in the *hospitalized* cohort it ranged from 6% (HealthVerity) to 28% (CU-AMC-DHC) and from 14% (SIDIAP) to 38% (CU-AMC-DHC), respectively.

**Table 1.**
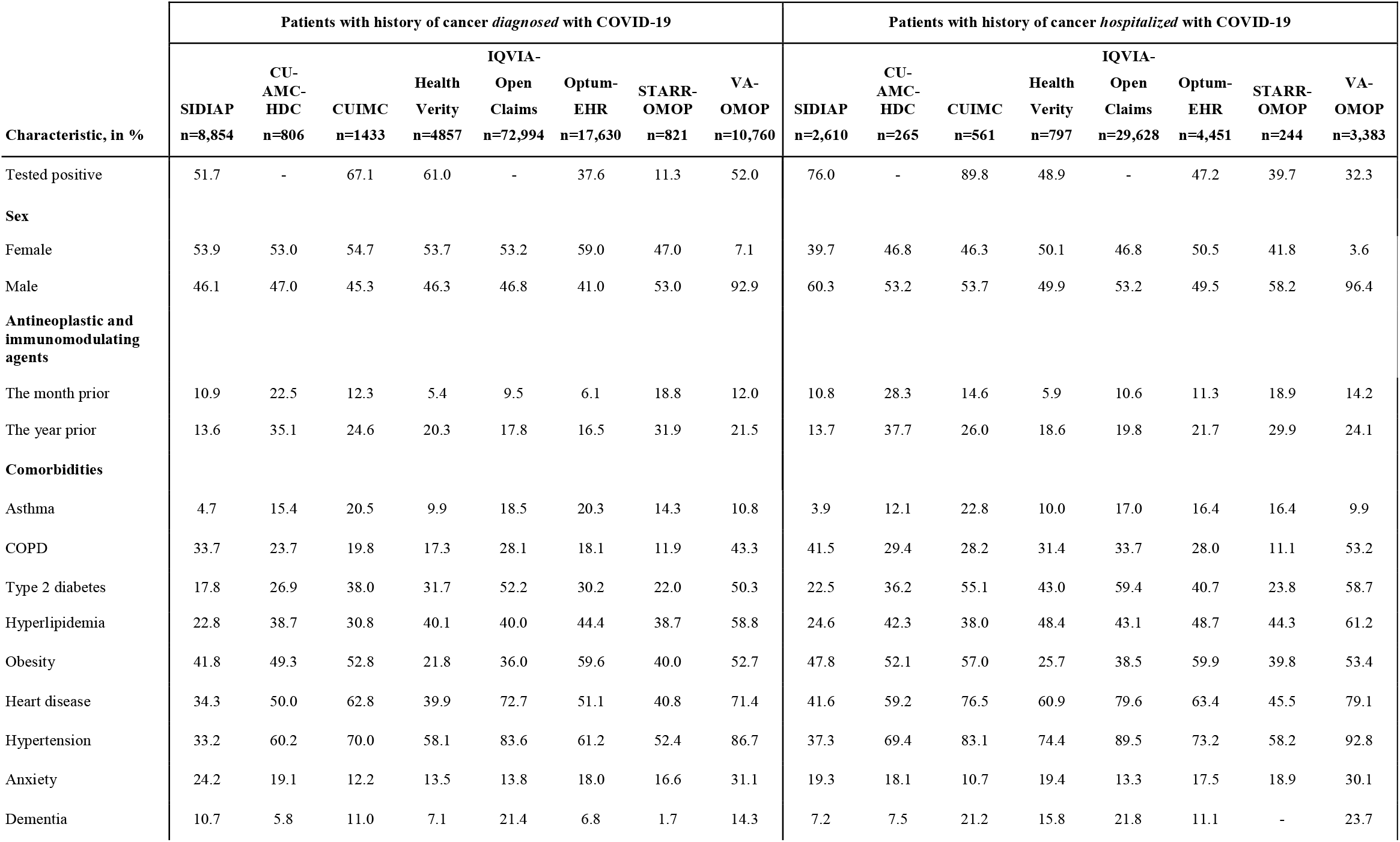
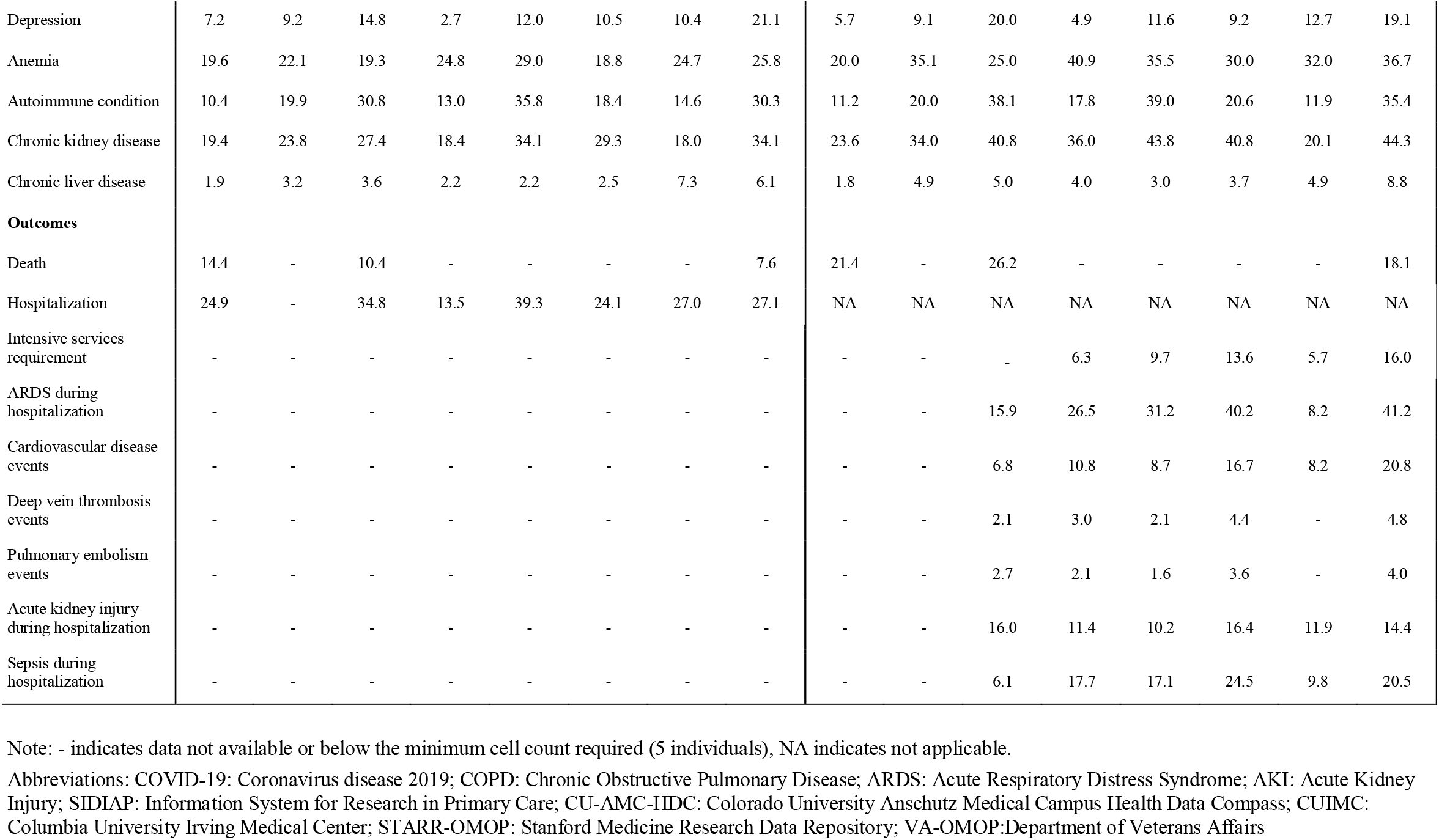
Demographics, comorbidities and outcomes among patients with a history of cancer *diagnosed* and *hospitalized* with COVID-19.

### Frequency of cancer types

For both COVID-19 cohorts, the frequency of the 26 selected cancer types can be consulted in Appendix Table A6. The top ten cancer types by frequency are reported in Table 2. In the COVID-19 *diagnosed* cohort, the most frequent cancers in four databases were breast (SIDIAP: 14.2%; CU-AMC-HDC: 7.3%; Optum-EHR: 6.7%; and STARR-OMOP: 12.3%) and prostate cancer (CUIMC: 6.1%; HealthVerity: 12.2%; IQVIA-OpenClaims: 7.1%; VA-OMOP: 18.1%). In all databases, non-Hodgkin’s lymphoma (NHL) was among the five most common cancers. Bladder, colorectal, leukemia, and lung cancer were also common (among the ten most frequent in at least 7 out of 8 databases).

**Table 2.** Top 10 cancer types among patients with a history of cancer *diagnosed* and *hospitalized* with COVID-19.

| Patients with a history of cancer *diagnosed* with COVID-19 |  |  |  |  |  |  |  |  |  |  |  |  |  |  |  |  |
| --- | --- | --- | --- | --- | --- | --- | --- | --- | --- | --- | --- | --- | --- | --- | --- | --- |
| Rank | SIDIAP<br>n=8,854 |  | CU-AMC-HDC<br>n=806 |  | CUIMC<br>n=1,433 |  | HealthVerity<br>n=4,857 |  | IQVIA-OpenClaims<br>n=72,994 |  | Optum-EHR<br>n=17,630 |  | STARR-OMOP<br>n=821 |  | VA-OMOP<br>n=10,760 |  |
|  | Cancer | % (95%CI) | Cancer | % (95%CI) | Cancer | % (95%CI) | Cancer | % (95%CI) | Cancer | % (95%CI) | Cancer | % (95%CI) | Cancer | % (95%CI) | Cancer | % (95%CI) |
| 1 | Breast | 14.2<br>(13.5-14.9) | Breast | 7.3<br>(5.5-9.1) | Prostate | 5.2<br>(4.0-6.4) | Prostate | 12.2<br>(11.3-13.1) | Prostate | 7.1<br>(6.9-7.3) | Breast | 6.7<br>(6.3-7.1) | Breast | 12.3<br>(10.0-14.6) | Prostate | 18.1<br>(17.4-18.8) |
| 2 | Colorrectal | 10.4<br>(9.8-11.0) | Prostate | 6.8<br>(5.1-8.5) | NHL | 5.4<br>(4.2-6.6) | Breast | 11.5<br>(10.6-12.4) | Breast | 6.0<br>(5.8-6.2) | Prostate | 4.9<br>(4.6-5.2) | Prostate | 10.2<br>(8.1-12.3) | Lung | 3.9<br>(3.5-4.3) |
| 3 | Prostate | 9.4<br>(8.8-10.0) | NHL | 5.0<br>(3.5-6.5) | Breast | 5.2<br>(4.0-6.4) | NHL | 4.6<br>(4.0-5.2) | NHL | 3.5<br>(3.4-3.6) | Uterus | 3.7<br>(3.4-4.0) | Liver | 7.7<br>(5.9-9.5) | NHL | 3.8<br>(3.4-4.2) |
| 4 | Bladder | 6.4<br>(5.9-6.9) | Lung | 3.8<br>(2.5-5.1) | Leukemia | 4.4<br>(3.3-5.5) | Colorrectal | 3.9<br>(3.4-4.4) | Lung | 2.7<br>(2.6-2.8) | Lip, oral<br>cavity and<br>pharynx | 3.7<br>(3.4-4.0) | Lung | 6.9<br>(5.2-8.6) | Bladder | 3.3<br>(3.0-3.6) |
| 5 | NHL | 3.0<br>(2.6-3.4) | Leukemia | 3.3<br>(2.1-4.5) | Liver | 3.3<br>(2.4-4.2) | Thyroid | 3.5<br>(3.0-4.0) | Leukemia | 2.6<br>(2.5-2.7) | NHL | 2.6<br>(2.4-2.8) | NHL | 5.8<br>(4.2-7.4) | Lip, oral<br>cavity and<br>pharynx | 2.9<br>(2.6-3.2) |
| 6 | Melanoma | 2.9<br>(2.6-3.2) | Melanoma | 3.0<br>(1.8-4.2) | Lung | 3.1<br>(2.2-4.0) | Leukemia | 2.8<br>(2.3-3.3) | Colorrectal | 2.6<br>(2.5-2.7) | Leukemia | 2.3<br>(2.1-2.5) | Thyroid | 5.2<br>(3.7-6.7) | Leukemia | 2.6<br>(2.3-2.9) |
| 7 | Leukemia | 2.7<br>(2.4-3.0) | Colorrectal | 2.9<br>(1.7-4.1) | Multiple<br>myeloma | 3.1<br>(2.2-4.0) | Lung | 2.7<br>(2.2-3.2) | Bladder | 1.7<br>(1.6-1.8) | Lung | 2.2<br>(2.0-2.4) | Lip, oral<br>cavity and<br>pharynx | 4.9<br>(3.4-6.4) | Colorrectal | 2.6<br>(2.3-2.9) |
| 8 | Uterus | 2.5<br>(2.2-2.8) | Multiple<br>myeloma | 2.5<br>(1.4-3.6) | Colorrectal | 2.7<br>(1.9-3.5) | Bladder | 2.5<br>(2.1-2.9) | Multiple<br>myeloma | 1.6<br>(1.5-1.7) | Colorrectal | 1.8<br>(1.6-2.0) | Leukemia | 3.8<br>(2.5-5.1) | Kidney | 2.5<br>(2.2-2.8) |
| 9 | Kidney | 2.4<br>(2.1-2.7) | Bladder | 2.2<br>(1.2-3.2) | Uterus | 2.0<br>(1.3-2.7) | Multiple<br>myeloma | 2.1<br>(1.7-2.5) | Liver | 1.5<br>(1.4-1.6) | Thyroid | 1.5<br>(1.3-1.7) | Colorrectal | 3.7<br>(2.4-5.0) | Liver | 1.8<br>(1.5-2.1) |
| 10 | Lip, oral<br>cavity and<br>pharynx | 1.4<br>(1.2-1.6) | Thyroid | 2.2<br>(1.2-3.2) | Kidney | 1.8<br>(1.1-2.5) | Kidney | 2.1<br>(1.7-2.5) | Kidney | 1.4<br>(1.3-1.5) | Bladder | 1.4<br>(1.2-1.6) | Bladder | 3.2<br>(2.0-4.4) | Larynx | 1.3<br>(1.1-1.5) |

**Patients with a history of cancer *hospitalized* with COVID-19**
| Rank | SIDIAP<br>n=2,610 |  | CU-AMC-HDC<br>n=265 |  | CUIMC<br>n=561 |  | HealthVerity<br>n=797 |  | IQVIA-OpenClaims<br>n=29,628 |  | Optum-EHR<br>n=4,451 |  | STARR-OMOP<br>n=244 |  | VA-OMOP<br>n=3,383 |  |
| --- | --- | --- | --- | --- | --- | --- | --- | --- | --- | --- | --- | --- | --- | --- | --- | --- |
|  | Cancer | %<br>(95%CI) | Cancer | %<br>(95%CI) | Cancer | %<br>(95%CI) | Cancer | %<br>(95%CI) | Cancer | %<br>(95%CI) | Cancer | %<br>(95%CI) | Cancer | %<br>(95%CI) | Cancer | %<br>(95%CI) |
| 1 | Prostate | 12.8<br>(11.5-14.1) | NHL | 6.4<br>(3.5-9.3) | Prostate | 7.7<br>(5.5-9.9) | Prostate | 10.8<br>(8.6-13.0) | Prostate | 8.5<br>(8.2-8.8) | Breast | 6.4<br>(5.7-7.1) | Prostate | 10.7<br>(6.8-14.6) | Prostate | 19.4<br>(18.1-20.7) |
| 2 | Colorectal | 11.9<br>(10.7-13.1) | Prostate | 6.4<br>(3.5-9.3) | NHL | 6.4<br>(4.4-8.4) | Breast | 9.2<br>(7.2-11.2) | Breast | 5.4<br>(5.1-5.7) | Prostate | 6.0<br>(5.3-6.7) | Lip, oral<br>cavity and<br>pharynx | 9.0<br>(5.4-12.6) | Lung | 5.7<br>(4.9-6.5) |
| 3 | Breast | 9.4<br>(8.3-10.5) | Lung | 6.0<br>(3.1-8.9) | Leukemia | 4.1<br>(2.5-5.7) | NHL | 7.4<br>(5.6-9.2) | NHL | 4.5<br>(4.3-4.7) | NHL | 4.2<br>(3.6-4.8) | Lung | 9.0<br>(5.4-12.6) | NHL | 4.6<br>(3.9-5.3) |
| 4 | Bladder | 8.5<br>(7.4-9.6) | Multiple<br>myeloma | 4.2<br>(1.8-6.6) | Lung | 3.7<br>(2.1-5.3) | Colorectal | 5.0<br>(3.5-6.5) | Lung | 4.0<br>(3.8-4.2) | Lung | 3.6<br>(3.1-4.1) | Breast | 7.4<br>(4.1-10.7) | Bladder | 3.9<br>(3.2-4.6) |
| 5 | NHL | 4.3<br>(3.5-5.1) | Leukemia | 4.2<br>(1.8-6.6) | Liver | 3.6<br>(2.1-5.1) | Leukemia | 4.1<br>(2.7-5.5) | Leukemia | 3.5<br>(3.3-3.7) | Lip, oral<br>cavity and<br>pharynx | 3.5<br>(3.0-4.0) | Liver | 6.6<br>(3.5-9.7) | Leukemia | 3.3<br>(2.7-3.9) |
| 6 | Leukemia | 4.2<br>(3.4-5.0) | Breast | 3.8<br>(1.5-6.1) | Colorectal | 3.4<br>(1.9-4.9) | Lung | 4.0<br>(2.6-5.4) | Colorectal | 3.0<br>(2.8-3.2) | Leukemia | 3.4<br>(2.9-3.9) | Thyroid | 6.1<br>(3.1-9.1) | Colorectal | 3.2<br>(2.6-3.8) |
| 7 | Kidney | 2.8<br>(2.2-3.4) | Liver | 3.8<br>(1.5-6.1) | Multiple<br>myeloma | 3.4<br>(1.9-4.9) | Multiple<br>myeloma | 3.4<br>(2.1-4.7) | Liver | 2.2<br>(2.0-2.4) | Uterus | 3.0<br>(2.5-3.5) | Pancreas | 5.3<br>(2.5-8.1) | Liver | 2.8<br>(2.2-3.4) |
| 8 | Melanoma | 2.3<br>(1.7-2.9) | - |  | Breast | 3.2<br>(1.7-4.7) | Bladder | 3.0<br>(1.8-4.2) | Multiple<br>myeloma | 2.2<br>(2.0-2.4) | Liver | 2.7<br>(2.2-3.2) | Leukemia | 4.9<br>(2.2-7.6) | Lip, oral<br>cavity and<br>pharynx | 2.8<br>(2.2-3.4) |
| 9 | Uterus | 1.8<br>(1.3-2.3) | - |  | Central<br>nervous<br>system | 2.0<br>(0.8-3.2) | Uterus | 2.8<br>(1.7-3.9) | Bladder | 2.0<br>(1.8-2.2) | Colorectal | 2.5<br>(2.0-3.0) | Oropharynx | 4.5<br>(1.9-7.1) | Kidney | 2.7<br>(2.2-3.2) |
| 10 | Liver | 1.5<br>(1.0-2.0) | - |  | Uterus | 1.8<br>(0.7-2.9) | Lip, oral<br>cavity and<br>pharynx | 2.5<br>(1.4-3.6) | Kidney | 1.7<br>(1.6-1.8) | Bladder | 2.4<br>(2.0-2.8) | - |  | Multiple<br>myeloma | 1.8<br>(1.4-2.2) |

Notes: - indicates data not available. A single individual can have multiple cancer types records.
Abbreviations: COVID-19: Coronavirus disease 2019; SIDIAP: Information System for Research in Primary Care; CU-AMC-HDC: Colorado University Anschutz Medical Campus Health Data Compass; CUIMC: Columbia University Irving Medical Center; STARR-OMOP: Stanford Medicine Research Data Repository; VA-OMOP: Department of Veterans Affairs; NHL: Non-Hodgkin's lymphoma

In the COVID-19 *hospitalized* cohort, prostate cancer was the most frequent cancer in all databases (ex aequo with NHL in CU-AMC-HDC, 6.4%), aside from Optum-EHR (breast cancer, 6.4%), in which it was the second most frequent. NHL was among the three most frequent cancers in all databases aside from SIDIAP and STARR-OMOP, where NHL was the fifth most common. Leukemia, liver and lung cancer were also within the top ten in the majority of databases. We did not observe meaningful differences (i.e. SMD >|0.1|) when comparing cancer types between the *diagnosed* and the *hospitalized* cohorts (Appendix Figure A3).

### Prior comorbidities

In both COVID-19 cohorts, the most common comorbidities were cardiometabolic conditions, which were more frequent in the US databases (especially VA-OMOP) than in the Spanish SIDIAP database. For example, hypertension ranged in the US from 52% to 87% (Spain: 32%) among *diagnosed* and from 58% to 93% (Spain: 33%) among *hospitalized* patients. Other common conditions include chronic obstructive pulmonary disease (COPD) (range in *diagnosed*: 12% to 43% (Spain: 34%), range in *hospitalized*: 11% to 53% (Spain: 42%)); and chronic kidney disease (range in *diagnosed*: 18% to 34% (Spain: 19%), range in *hospitalized*: 20% to 44% (Spain: 24%)). The prevalence of the full range of prior conditions summarized is shown in Figure 2. Several comorbidities were more frequent among patients *hospitalized* compared to patients *diagnosed* (SMD>0.1): heart disease (all databases except STARR-OMOP) and chronic kidney disease, hypertension and type 2 diabetes (all except SIDIAP and STARR-OMOP) (Figure 3).

**Figure 1.**
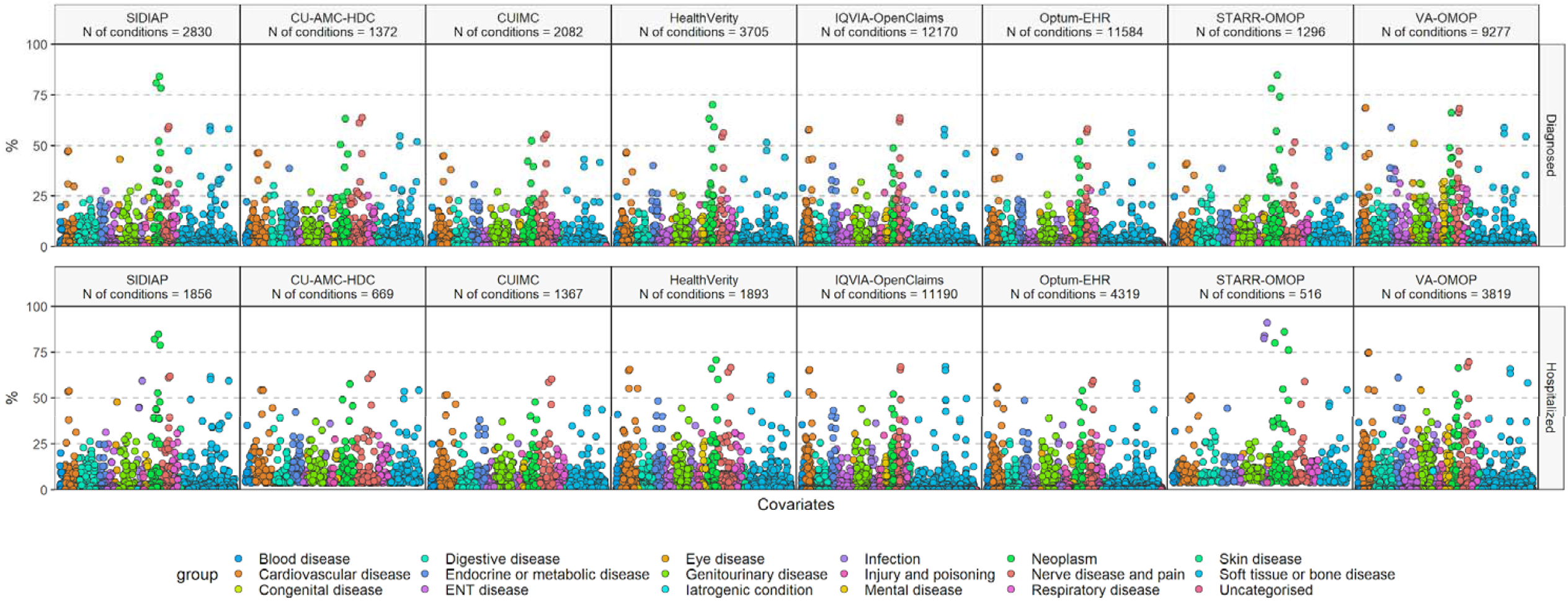
Prevalence of baseline conditions among patients with a history of cancer *diagnosed* and *hospitalized* with COVID-19. Each dot represents one of these conditions with the colour indicating the type of condition. Notes: only conditions meeting the minimum count requirement (5 individuals) are shown. Abbreviations: COVID-19: Coronavirus disease 2019; SIDIAP: Information System for Research in Primary Care; CU-AMC-HDC: Colorado University Anschutz Medical Campus Health Data Compass; CUIMC: Columbia University Irving Medical Center; STARR-OMOP: Stanford Medicine Research Data Repository; VA-OMOP: Department of Veterans Affairs.

**Figure 2.**
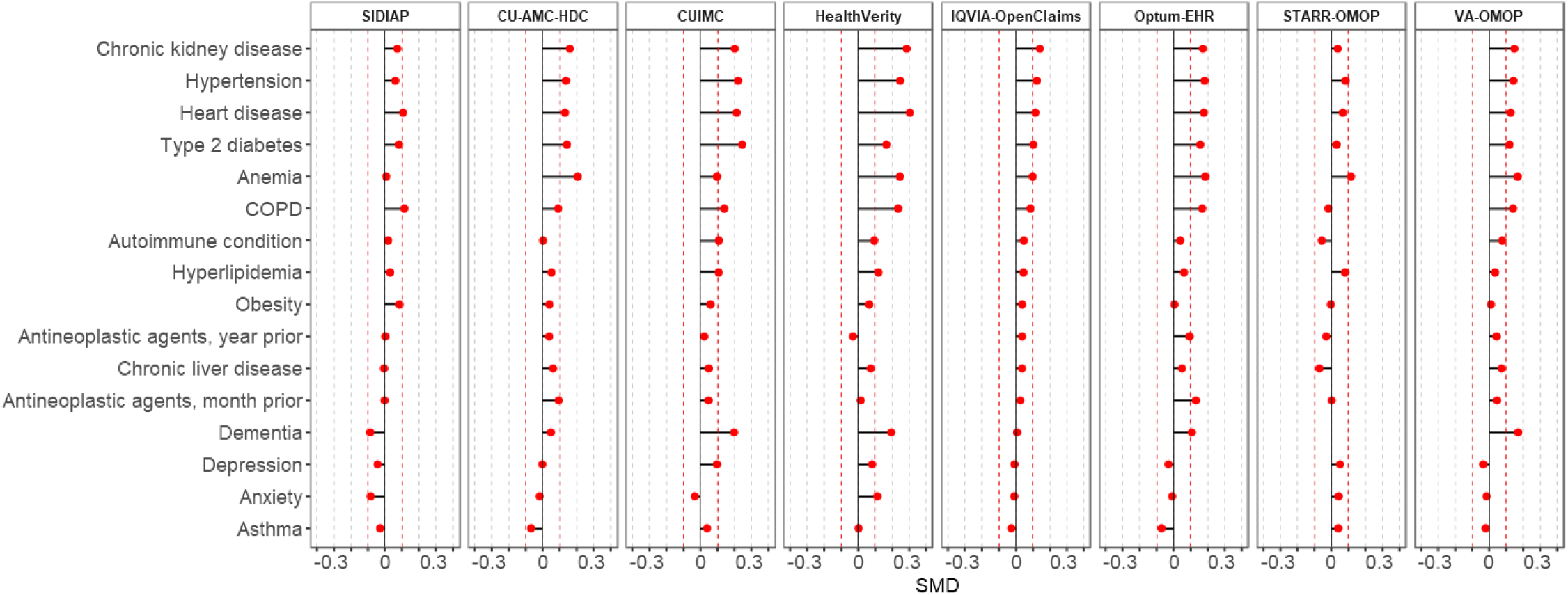
Standardized mean differences of selected baseline conditions between patients with a history of cancer *diagnosed* and *hospitalized* with COVID-19. SMD<0 indicates that the prevalence was greater in patients *diagnosed*, SMD>0 indicates that the prevalence was greater in patients *hospitalized*. Notes: SMD were calculated for cancer types meeting the minimum count required (5 individuals) in each database and cohort. Cancer types ordered according to SMD descending values in the largest database (IQVIA-OpenClaims). Red-dotted lines indicate a SMD of |0.1|. Abbreviations: COVID-19: Coronavirus disease 2019; COPD: Chronic Obstructive Pulmonary Disease; ARDS: Acute Respiratory Distress Syndrome; AKI: Acute Kidney Injury; CU-AMC-HDC: Colorado University Anschutz Medical Campus Health Data Compass; CUIMC: Columbia University Irving Medical Center; STARR-OMOP: Stanford Medicine Research Data Repository; VA-OMOP:Department of Veterans Affairs; SMD: Standardized Mean Difference

**Figure 3.**
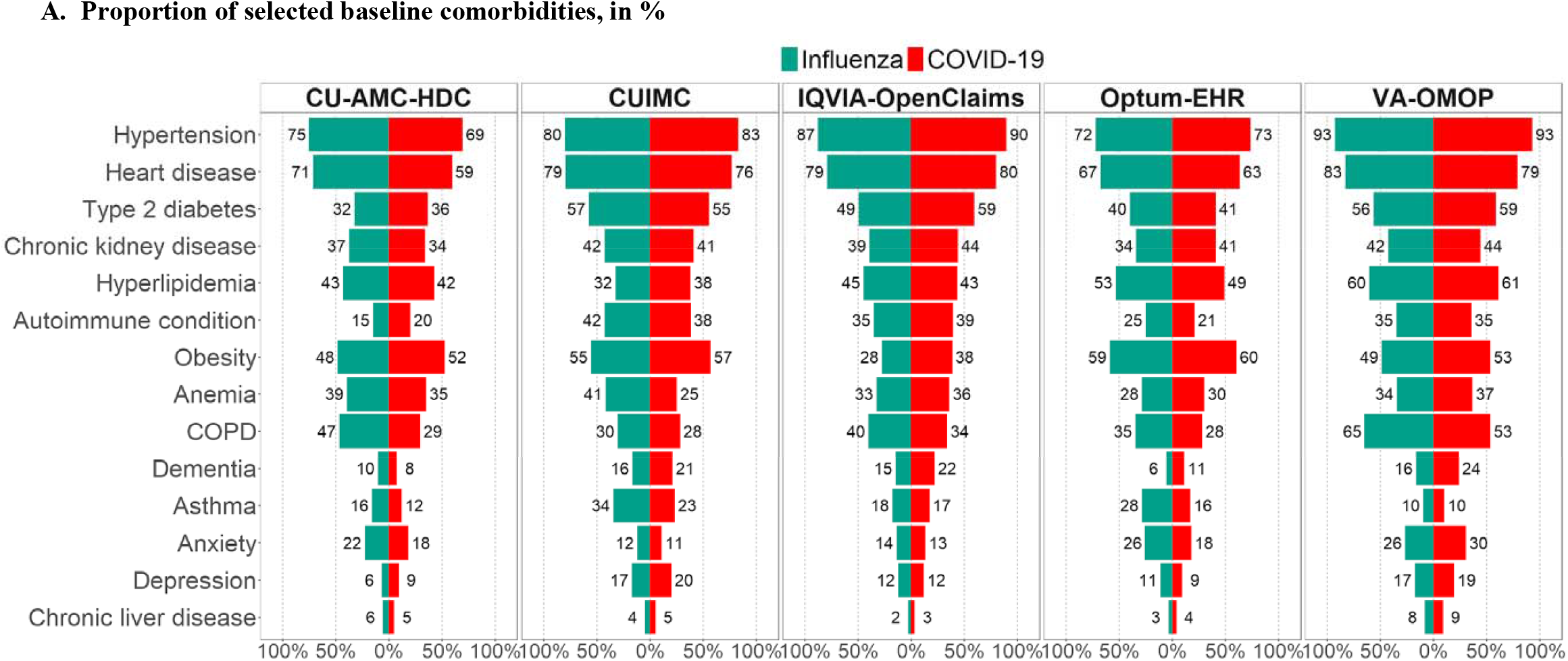
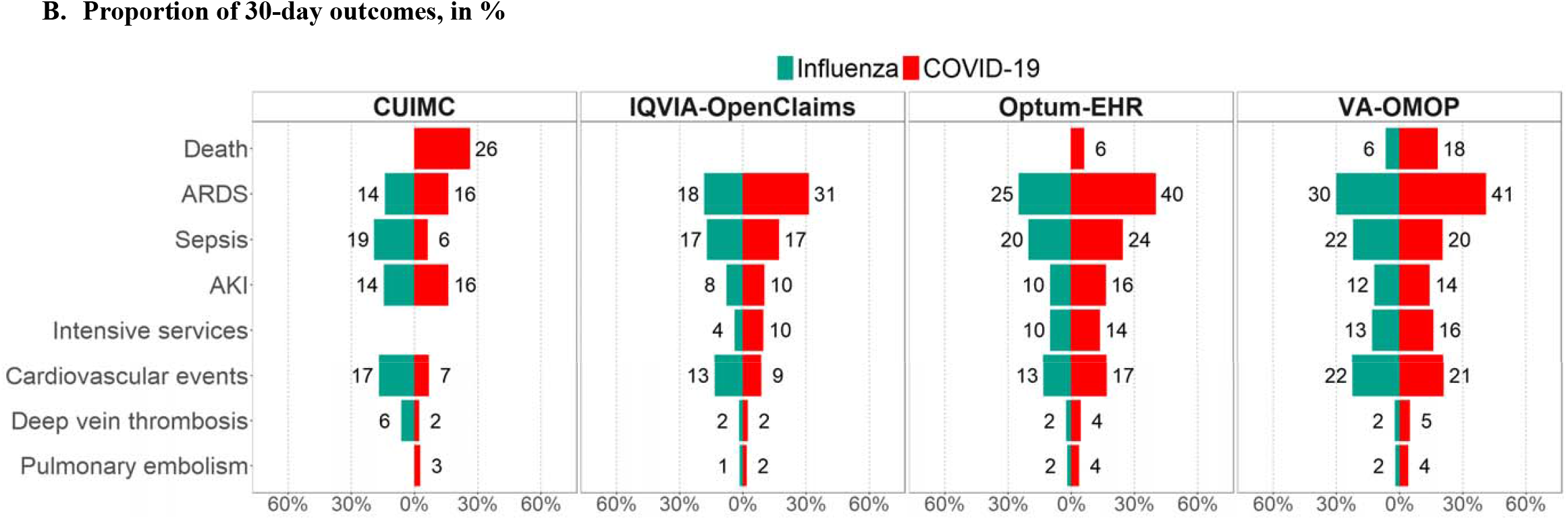
Baseline comorbidities and 30-day outcomes among patients with a history of cancer *hospitalized* with COVID-19 and with seasonal influenza. Notes: Comorbidities and outcomes are shown if meeting the minimum count required (5 individuals) in each database and cohort. Comorbidities and outcomes ordered according to descending values in the largest database (IQVIA-OpenClaims). Abbreviations: COVID-19: Coronavirus disease 2019; COPD: Chronic Obstructive Pulmonary Disease; ARDS: Acute Respiratory Distress Syndrome; AKI: Acute Kidney Injury; CU-AMC-HDC: Colorado University Anschutz Medical Campus Health Data Compass; CUIMC: Columbia University Irving Medical Center; STARR-OMOP: Stanford Medicine Research Data Repository; VA-OMOP:Department of Veterans Affairs

**Figure 4.**
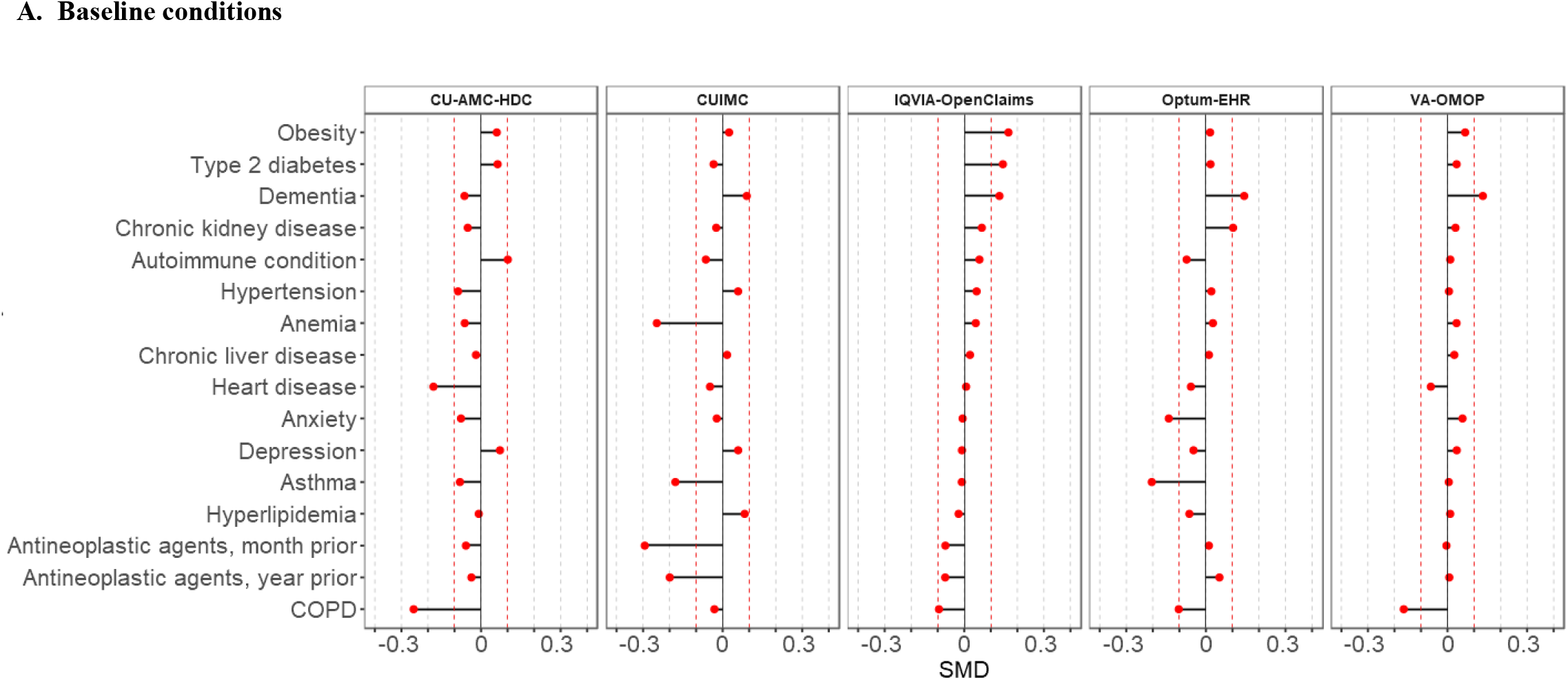
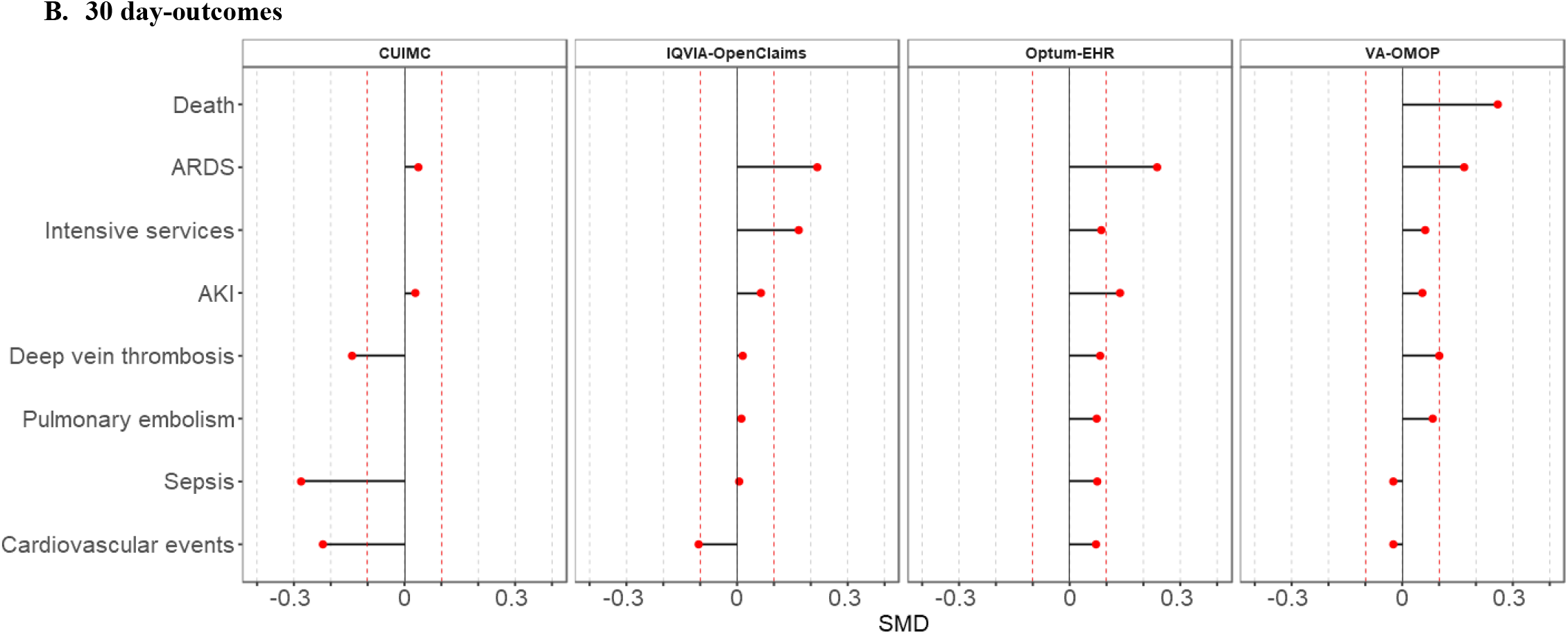
Standardized mean differences of selected baseline conditions and 30-day outcomes between patients with a history of cancer *hospitalized* with COVID-19 and with seasonal influenza. SMD <0 indicates that the prevalence was greater in patients with seasonal influenza, SMD>0 indicates that the prevalence was greater in patients hospitalized. Notes: SMD were calculated for conditions and outcomes meeting the minimum count required (5 individuals) in each database and cohort..Comorbidities and outcomes ordered according to SMD descending values in the largest database (IQVIA-OpenClaims). Red-dotted lines indicate a SMD of |0.1|. Abbreviations: COVID-19: Coronavirus disease 2019; COPD: Chronic Obstructive Pulmonary Disease; ARDS: Acute Respiratory Distress Syndrome; AKI: Acute Kidney Injury; CU-AMC-HDC: Colorado University Anschutz Medical Campus Health Data Compass; CUIMC: Columbia University Irving Medical Center; STARR-OMOP: Stanford Medicine Research Data Repository; VA-OMOP:Department of Veterans Affairs; SMD: Standardized Mean Difference

### Thirty-day outcomes

In the COVID-19 *diagnosed* cohort, hospitalization rates in the US databases ranged from 14% to 40% (Spain: 25%) and occurrence of death from 8% to 10% (Spain: 14%).

In the COVID-19 *hospitalized* cohort, outcomes were heterogeneous across databases, occurring less frequently in STARR-OMOP and more frequently in VA-OMOP. ARDS ranged from 8% to 41%, although it was higher than 30% in 3 out of 6 databases with data available (IQVIA-OpenClaims, Optum-EHR, VA-OMOP). Sepsis was also common, ranging from 6% to 25%. Cardiovascular disease events and AKI ranged from 7% to 21% and from 10% to 16%, respectively. Thromboembolic events were less frequent, with deep vein thrombosis ranging from 2% to 5% and pulmonary embolism from 2% to 4%. Intensive services requirement ranged from 6% to 16%, whereas occurrence of death ranged from 18% to 26% in the US (Spain: 21%).

### Comparison of patients with a history of cancer hospitalized with COVID-19 to those hospitalized with seasonal influenza

The characteristics of patients with a history of cancer hospitalized with seasonal influenza are reported in Appendix Table A8. Overall, these patients were predominantly male and the majority clustered around the ages of 60 to 85 years old (Appendix Figure A5). When comparing the frequency of cancer types between COVID-19 and influenza patients, we did not observe consistent differences across databases (Appendix Figure A4). The distribution of comorbidities was similar in both groups, with few exceptions (Figure 2A). For example, COPD was more common among patients with influenza in 3 databases: CU-AMC-HDC, Optum-EHR and VA-OMOP (Figure 3A). Aside from CUIMC, outcomes were more common in the COVID-19 cohort, especially ARDS, ranging from 16% to 41% in the COVID-19 cohort and from 14% to 30% in the influenza cohort (SMD>0.2 in IQVIA-OpenClaims and Optum EHR, SMD>0.1 in VA-OMOP). Occurrence of death was higher among COVID-19 patients compared to influenza patients in VA-OMOP: 18% vs 6 (SMD>0.2) (Figures 2B and 3B).

## Discussion

In this multinational cohort study, we described in detail the characteristics of 118,155 patients with a history of cancer and COVID-19, including outcomes that have been rarely reported in this population. The lifetime cancer prevalence was higher among those *hospitalized* compared to all those *diagnosed* with COVID-19. In both COVID-19 cohorts of patients with history of cancer, the most frequent cancer type was either prostate or breast cancer; hematological malignancies were also frequent. Comorbidities were common in both cohorts but were higher among those *hospitalized*. Occurence of death ranged from 7% to 14% among those *diagnosed* and from 18% to 26% among those *hospitalized*. When compared to patients with a history of cancer hospitalized with seasonal influenza, patients hospitalized with COVID-19 had a similar distribution of sex, age and comorbidities but had more severe outcomes.

According to recent estimates, the lifetime cancer prevalence in the US is 5%.^19^ In Spain, the 5-year cancer prevalence is 1.7% (data on the lifetime cancer prevalence is unavailable to our knowledge).^16^ Although our results were heterogeneous, we found that the lifetime cancer prevalence was higher among COVID-19 cases compared to the general population, ranging from 4% to 25% in the *diagnosed* and from 11% to 40% in the *hospitalized* cohort. Even though comparisons are limited due to differences in the definition of cancer, these prevalences are also higher than prior reports on COVID-19 patients at hospital settings. In early studies from China, less than 3% of inpatients with COVID-19 had cancer, whereas studies from Europe and the US have reported a cancer prevalence of 6% to 11%.^20–25^ A Danish study, however, found a lifetime cancer prevalence among patients hospitalized with COVID-19 of 17%, which is in line with our results.^6^ Further, the lifetime cancer prevalence was consistently higher in patients *hospitalized* compared to those *diagnosed*, which might partially be due to the older age of the former.

The most lifetime-prevalent cancer types in the US population are prostate (in men) and breast (in women) cancer.^19^ Two hematological malignancies are also frequent among cancer survivors: NHL is the fifth/sixth more frequent (men and women, respectively), whereas leukemia is the ninth in men. We found that prostate and breast cancer were the most frequent cancers in COVID-19 patients, which is in line with the general cancer population. These cancer types were similarly common among *diagnosed* patients (where females predominated slightly), whereas in *hospitalized* patients (where males were the majority), prostate cancer was more frequent than breast cancer. On the other hand, hematological malignancies were more common than expected in our cohorts (both COVID-19 and influenza). For example, in the COVID-19 *hospitalized* cohorts, NHL, leukemia and multiple myeloma were among the third, fifth, and tenth more common cancers, respectively. Prior studies have reported an overrepresentation of hematological malignancies in COVID-19 patients compared to the general population.^5,26^ This is of particular concern in light of evidence suggesting that patients with hematological malignancies have an increased risk of COVID-19 mortality compared to those with solid cancers.^5^ The fact that hematological malignancies were more common than expected in both the *diagnosed* and the *hospitalized* COVID-19 cohorts raises questions on whether patients with these malignancies are more exposed or more vulnerable to SARS-CoV-2 infection, or both. Because of the underlying severity of their disease, patients with hematological malignancies are particularly at risk of HAI ^27^ and SARS-CoV-2 outbreaks have already been described in hematological units.^28^

As expected, patients with a history of cancer were older and had more comorbidities than overall COVID-19 cases. A meta-analysis comprising 12,149 COVID-19 cases (mostly hospitalized) found hypertension (23%), heart failure (20%) and diabetes (12%) were the most common comorbidities.^29^ These numbers are substantially lower than what we have observed. Similarly, a study characterizing COVID-19 inpatients found a prevalence of heart disease of 27% in SIDIAP (vs 42% in this study), 29% in CUIMC (vs 77%) and 48% in VA-OMOP (vs 80%).^14^ Compared to studies reporting comorbidities among patients with cancer, we also found higher prevalences of comorbidities. For example, chronic kidney disease (range 20-44%), diabetes (23-59%) and obesity (26-60%) were higher in our *hospitalized* cohort than in a study including COVID-19 inpatients with a history of solid cancer (16%, 22% and 10% had chronic kidney disease, diabetes and obesity, respectively).^23^ In addition, we observed that heart disease, chronic kidney disease and type 2 diabetes were meaningfully higher among those *hospitalized* compared to those *diagnosed*. These conditions have been previously reported as potential risk factors for hospitalization, increased severity and mortality among COVID-19 cases.^30^ The role of comorbidities should be taken into consideration when designing future studies assessing the effect of cancer on COVID-19-related health outcomes, as failing to adjust for some comorbidities or adjusting for others (over-adjustment) could lead to confounding and/or selection bias given the high comorbidity burden.

According to the World Health Organization, in June 2020 the case-fatality ratio among confirmed COVID-19 cases was 11.4% in Spain and 5.0% in the US,^31^ which is lower than the all-cause mortality observed both in the *diagnosed* (Spain: 14%, US: 8-10%) and *hospitalized* (Spain: 21%, US: 18-26%) cohorts. A meta-analysis including 18,650 patients with cancer and COVID-19 (mostly from the hospital setting) reported a pooled case mortality rate of 25.6% (95% CI: 22.0-29.5%),^32^ in line with our findings in the *hospitalized* cohort. Undoubtedly, increased age and underlying comorbidities play a substantial role in COVID-19 related mortality among these patients.

Finally, we have compared patients with cancer history *hospitalized* with COVID-19 to those with seasonal influenza as a benchmark. We previously showed that COVID-19 patients are more often male, younger and less likely to have respiratory and cardiovascular diseases than influenza patients.^14^ Interestingly, we have observed that COVID-19 and influenza patients with a history of cancer have a similar sex and age distribution and are of comparable health status. Despite this similarity, patients with cancer history and COVID-19 had worse outcomes than those with cancer history and influenza, including mortality, intensive services requirements, ARDS and thromboembolic events, which might be expected given the virulence of the disease.

This study has many major strengths, such as its large size. We have reported more than 10,000 aggregate characteristics from more than 100,000 patients from eight different databases and two countries. The diverse healthcare settings and populations described in this study, together with our multinational approach, increases the generalizability of our findings. Further, as CHARYBDIS is an ongoing study, we expect that more databases from a range of countries will provide sufficient data on the cancer population as the pandemic evolves. By including only individuals with at least one year of observation time available prior to index date, we have comprehensively captured baseline comorbidities, which could explain the higher prevalence of selected comorbidities in our cohorts compared to prior studies. In addition, we have ensured confidentiality throughout the study using a federated analysis approach. Finally, for the purposes of transparency and reproducibility, our methods, tools, and results are all publicly available.

However, this study also has several limitations. First, we were not able to provide detailed cancer information, such as the year of cancer diagnosis or stage of tumour at diagnosis; nor identify patients with active cancer treatment, and we were not able to characterize patients stratified by cancer types due to small sample sizes. Secondly, although we included patients with a clinical COVID-19 diagnosis to reduce selection bias due to testing restrictions during the first months of the pandemic, we cannot exclude that we have incurred some false positives. Additionally, we did not have information on the cause of death and reported all-cause death as an outcome. Third, the differences found in the COVID-19/seasonal influenza comparison may have been influenced by temporal changes in clinical practice standards and coding. Further, the use of influenza vaccination among high-risk population groups likely contributed to the observed low proportion of adverse events among influenza patients.

Finally, we found heterogeneous results across databases, which hinders the interpretation of our findings. Heterogeneity across data sources is a known phenomenon when using real-world data that reflects the existence of different coding practices, observation period, healthcare settings and populations. Indeed, the fact that the percentages of several cancer types were very low in some databases might be explained by differences in coding practices across data sources (i.e. some codes might be more widely used in some databases than others). Similarly, the use of routinely-collected data could have led to an underestimation of the lifetime cancer prevalence, cancer types, comorbidities, and outcomes due to incomplete reporting. While the interpretation of heterogeneous results is challenging, these also provide valuable insights into the particularities of each each setting, which is the objective of generalizability of real world data for certain populations. Yet, despite this heterogeneity, we found consistent patterns when comparing characteristics across cohorts, which lends credence to our results.

## Conclusion

This in-depth characterization revealed that COVID-19 patients with a history of cancer are mostly aged above 65 years old and have a myriad of comorbidities that may explain the high frequency of severe COVID-19-outcomes observed in this population. In addition, we found that hematological malignancies were more frequent than expected. These findings are foundational for guiding future studies and highlight the importance of protecting patients with cancer while guaranteeing cancer care continuity during the pandemic.

## Data Availability

Open Science is a guiding principle within OHDSI.  As such, we provide unfettered access to all open-source analysis tools employed in this study via https://github.com/ohdsi-studies/Covid19CharacterizationCharybdis, as well as all data and results artefacts that do not include patient-level health information via https://data.ohdsi.org/Covid19CharacterizationCharybdis/. Data partners contributing to this study remain custodians of their individual patient-level health information and hold either IRB exemption or approval for participation.

## Abbreviations

ARDS: Acute Respiratory Distress Syndrome;
CDM: Common Data Model;
CHARYBDIS: Characterizing Health Associated Risks, and Your Baseline Disease In SARS-COV-2;
CI: Confidence Interval;
COPD: Chronic Obstructive Pulmonary Disease:
COVID-19: Coronavirus disease 2019;
CU-AMC-DHC: Colorado University Anschutz Medical Campus Health Data Compass;
CUIMC: Columbia University Irving Medical Center;
EHR: Electronic Health Record;
EHR: Electronic Health Record;
IRB: Institutional Review Board;
OHDSI: Observational Health Data Sciences and Informatics;
HAI: Healthcare-associated infection;
OMOP: Observational Medical Outcomes Partnership;
NHL: Non-Hodgkin’s lymphoma;
SARS-CoV-2: Severe Acute respiratory Syndrome Coronavirus-2;
SIDIAP: Information System for Research in Primary Care;
SMD: Standardized Mean Differences;
SNOMED: Systematized Nomenclature of Medicine;
STARR-OMOP: Stanford Medicine Research Data Repository;
UK: United Kingdom;
US: United States;
VA-OMOP: United States Department of Veterans Affairs.

## Acknowledgements

We would like to acknowledge the patients who suffered from or died of this devastating disease, and their families and carers. We would also like to thank the healthcare professionals involved in the management of COVID-19 during these challenging times, from primary care to intensive care units.

## Ethical approval

All the data partners received Institutional Review Board (IRB) approval or exemption. STARR-OMOP had approval from IRB Panel #8 (RB-53248) registered to Leland Stanford Junior University under the Stanford Human Research Protection Program (HRPP). The research was approved by the Columbia University Institutional Review Board as an OHDSI network study. SIDIAP analyses were approved by the Clinical Research Ethics Committee of the IDIAPJGol (project code: 20/070-PCV). Approval for CPRD was provided by the Independent Scientific Advisory Committee (ISAC). For IQVIA-OpenClaims, approval is provided for OHDSI community studies. This is a retrospective database study on de-identified data and it is deemed not human subject research.

## Transparency declaration

Lead authors affirm that the manuscript is an honest, accurate, and transparent account of the study being reported; that no important aspects of the study have been omitted; and that any discrepancies from the study as planned have been explained.

## Contributorship statement

ER, APU, PR, DPA, KK and TDS conceived and designed the study. MA, CB, WC, FD, SLD, TF, GH, KL, MEM, KJ, AO, JP, CR, LMS, NS, MS and TDS coordinated data contributions at their respective sites. AP, AGS, SFB, JDP, KK and TDS analyzed the data. ER and AP produced the figures and tables. ER, MR, AG, LH, DRM, FN, IS, KK and TDS interpreted the data. ER and TDS searched the literature and wrote the first draft with insightful contributions from MR, DP, AG, LH, DRM, FN, APU, LMS, IS, VS, KK and TDS. All authors contributed to the revision of the first draft, reviewed and approved the final version of the manuscript.

## Data Sharing

Open Science is a guiding principle within OHDSI.□ As such, we provide unfettered access to all open-source analysis tools employed in this study via https://github.com/ohdsi-studies/Covid19CharacterizationCharybdis, as well as all data and results artefacts that do not include patient-level health information via https://data.ohdsi.org/Covid19CharacterizationCharybdis/. Data partners contributing to this study remain custodians of their individual patient-level health information and hold either IRB exemption or approval for participation.

