## Appendix for "Characteristics and outcomes of 118,155 COVID-19 individuals with a history of cancer in the United States and Spain"

Talita Duarte-Salles

Fundació Institut Universitari per a la recerca a l'Atenció Primària de Salut Jordi Gol i Gurina (IDIAPJGol)

Gran Via Corts Catalanes, 587 àtic

08007 Barcelona - Spain

**Appendix**

#### Appendix Figure A1. Selection process for inclusion in this study of databases contributing to CHARYBDIS

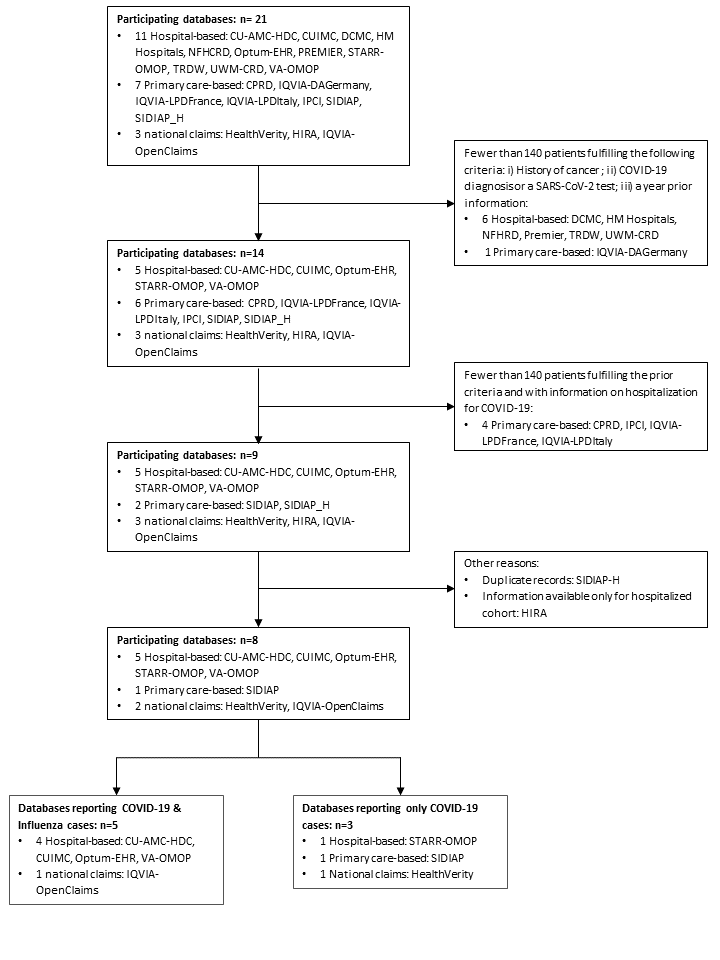

Abbreviations: CHARYBDIS: Characterizing Health Associated Risks and Your Baseline Disease In SARS-COV-2; CU-AMC-HDC: Colorado University Anschutz Medical Campus Health Data Compass; CUIMC:Columbia University Irving Medical Center ; DCMC: Daegu Catholic University Medical Center; NFHCRD: Nanfang Hospital COVID-19 Research Database; Optum-EHR: Optum® de-identified COVID-19 Electronic Health Record Dataset; STARR-OMOP: Stanford Medicine Research Data Repository; TRDW: Tufts Research Data Warehouse; UWM-CRD: UW Medicine COVID Research Dataset; VA-OMOP: Veterans Affairs; CPRD: Clinical Practice Research Datalink; IQVIA-LPDFrance: IQVIA-Longitudinal Patient Data France; IQVIA-LPDItaly: IQVIA-Longitudinal Patient Data Italy; IPCI: Integrated Primary Care Information; SIDIAP: Information System for Research in Primary Care; SIDIAP– H: SIDIAP-Hospitalization Linked Data; HIRA: Health Insurance Review & Assessment Service.

#### Appendix Table A1. Description of the databases from CHARYBDIS included in this study

| **Institution Name** | **Database** | **Database Description** | **Country** |
| --- | --- | --- | --- |
| IDIAPJGol | The Information System for Research in Primary Care (SIDIAP) | The Information System for Research in Primary Care (SIDIAP; www.sidiap.org) is a primary care records database that covers approximately 80% of the population of Catalonia; North-East Spain. Healthcare is universal and tax-payer funded in the region; and primary care physicians are gatekeepers for all care and responsible for repeat prescriptions. | Spain |
| University of Colorado | Colorado University Anschutz Medical Campus Health Data Compass (CU-AMC-HDC) | Health Data Compass (HDC) is a multi-institutional data warehouse. HDC contains inpatient and outpatient electronic medical data including patient; encounter; diagnosis; procedures; medications; laboratory results from two electronic medical record systems (UCHealth and Children's Hospital of Colorado); state-level all-payers claims data; and the Colorado death registry.. Acknowledgement statement: Supported by the Health Data Compass Data Warehouse project (healthdatacompass.org). | United States |
| Columbia | Columbia University Irving Medical Center (CUIMC) | The clinical data warehouse of New York-Presbyterian Hospital/Columbia University Irving Medical Center; New York; NY; based on its current and previous electronic health record systems; with data spanning over 30 years and including over 6 million patients | United States |
| Health Verity | HealthVerity | This HealthVerity derived data set contains de-identified patient information with an antibody and/or diagnostic test for COVID-19 linked to all available Medical Claims and Pharmacy Data from select private data providers participating in the HealthVerity marketplace. | United States |
| IQVIA | IQVIA-OpenClaims | Pre-adjudicated claims covering over 300 Million lives (~80% of the US) collected from office-based physicians and specialists via office management software and clearinghouse switch sources for the purpose of reimbursement. | United States |
| Optum | Optum® de-identified COVID-19 Electronic Health Record Dataset (Optum-EHR) | Optum de-identified Electronic Health Record Dataset represents Humedica Electronic Health Record data; a medical records database for patients receiving a COVID-19 diagnosis record or lab test for SARS-CoV-2. The medical record data includes clinical information; inclusive of prescriptions as prescribed and administered; lab results; vital signs; body measurements; diagnoses; procedures; and information derived from clinical Notes using Natural Language Processing (NLP). | United States |
| Stanford | Stanford Medicine Research Data Repository (STARR-OMOP) | A clinical data warehouse containing live Epic data from Stanford Health Care; the Stanford Children’s Hospital; the University Healthcare Alliance and Packard Children's Health Alliance clinics.  Reference: Datta S; Posada J; Olson G; et al. A new paradigm for accelerating clinical data science at Stanford Medicine. arXiv 2020; published online March 17. http://arxiv.org/abs/2003.10534 (accessed Aug 20; 2020). | United States |
| Department of Veterans Affairs | VA-OMOP | VA-OMOP data reflects the national Department of Veterans Affairs health care system; which is the largest integrated provider of medical and mental health services in the United States. Care is provided at 170 VA Medical Centers and 1;063 outpatient sites serving more than 9 million enrolled Veterans each year. | United States |

#### Appendix Table A2. Definitions and codes used to identify patients with COVID-19 and seasonal influenza; patients with a history of cancer and comorbidities at baseline and 30-day outcomes

| **Name** | **Definition** | **Included Codes** |
| --- | --- | --- |
| **Cohorts** |  |  |
| Persons with a COVID-19 diagnosis record or a SARS-CoV-2 positive test with at least 365d prior observation | Initial Event Cohort  People having any of the following:   - a measurement of SARS-CoV-2 positive test measurement pre-coordinated   - occurrence start is after 2019-12-01 - a measurement of SARS-CoV-2 test measurement   - occurrence start is after 2019-12-01   - value as concept is any of: Detected; Detected; Positive; Positive; Present; Present - an observation of SARS-CoV-2 test measurement   - occurrence start is after 2019-12-01   - value as concept is any of: Detected; Detected; Positive; Positive; Present; Present - a condition occurrence of COVID-19 conditions   - occurrence start is after 2019-12-01   with continuous observation of at least 0 days prior and 0 days after event index date; and limit initial events to: earliest event per person.  Inclusion Rules  Inclusion Criteria #1: has >=365 prior observation  Having all of the following criteria:   - at least 1 occurrences of an observation period - where event starts between all days Before and 365 days Before index start date and event ends between 0 days Before and all days After index start date | <https://atlas.ohdsi.org/#/cohortdefinition/200> |
| Persons hospitalized with a COVID-19 diagnosis record or a SARS-CoV-2 positive test with at least 365d prior observation | People having any of the following:   - a visit occurrence of Inpatient Visit   - occurrence start is after 2019-12-01   with continuous observation of at least 0 days prior and 0 days after event index date; and limit initial events to: all events per person.  For people matching the Primary Events; include:  Having any of the following criteria:   - at least 1 occurrences of a measurement of SARS-CoV-2 positive test measurement pre-coordinated   where event starts between 21 days Before and all days After index start date and event starts between all days Before and 0 days After index end date or at least 1 occurrences of a measurement of SARS-CoV-2 test measurement   - value as concept is any of: Detected; Detected; Positive; Positive; Present; Present - where event starts between 21 days Before and all days After index start date and event starts between all days Before and 0 days After index end date - or at least 1 occurrences of an observation of SARS-CoV-2 test measurement^4^   - value as concept is any of: Detected; Detected; Positive; Positive; Present; Present - where event starts between 21 days Before and all days After index start date and event starts between all days Before and 0 days After index end date - or at least 1 occurrences of a condition occurrence of COVID-19 conditions^1^ - where event starts between 21 days Before and all days After index start date and event starts between all days Before and 0 days After index end date   Limit cohort of initial events to: earliest event per person.  Inclusion Rules  Inclusion Criteria #1: has >=365 prior observation  Having all of the following criteria:   - at least 1 occurrences of an observation period - where event starts between all days Before and 365 days Before index start date and event ends between 0 days Before and all days After index start date | <https://atlas.ohdsi.org/#/cohortdefinition/197> |
| Persons hospitalized with influenza diagnosis or positive test 2017-2018 with at least 365d prior observation | People having any of the following:   - a visit occurrence of Inpatient Visit   - occurrence start is between 2017-09-01 and 2018-04-01 (inclusive)   with continuous observation of at least 0 days prior and 0 days after event index date; and limit initial events to: all events per person.  For people matching the Primary Events; include:  Having any of the following criteria:   - at least 1 occurrences of a condition occurrence of Influenza conditions - where event starts between 21 days Before and all days After index start date and event starts between all days Before and 0 days After index end date - or at least 1 occurrences of a measurement of Influenza positive measurements pre-coordinated - where event starts between 21 days Before and all days After index start date and event starts between all days Before and 0 days After index end date - or at least 1 occurrences of a measurement of Influenza test measurements excluding antibody testing   - value as concept is any of: Detected; Detected; Positive; Present; Present; Positive - where event starts between 21 days Before and all days After index start date and event starts between all days Before and 0 days After index end date - or at least 1 occurrences of an observation of Influenza test measurements excluding antibody testing   - value as concept is any of: Detected; Detected; Positive; Positive; Present; Present - where event starts between 21 days Before and all days After index start date and event starts between all days Before and 0 days After index end date   Limit cohort of initial events to: all events per person.  Inclusion Rules  Inclusion Criteria #1: has >=365d of prior observation  Having all of the following criteria:   - at least 1 occurrences of an observation period - where event starts between all days Before and 365 days Before index start date and event ends between 0 days Before and all days After index end date   Inclusion Criteria #2: does not have hospitalization for influenza in the 6 months preceding admission  Having all of the following criteria:   - exactly 0 occurrences of a visit occurrence of Inpatient Visit   - Having any of the following criteria:     - at least 1 occurrences of a condition occurrence of Influenza conditions     - where event starts between 21 days Before and all days After index start date and event starts between all days Before and 0 days After index end date     - or at least 1 occurrences of a measurement of Influenza positive measurements pre-coordinated     - where event starts between 21 days Before and all days After index start date and event starts between all days Before and 0 days After index end date     - or at least 1 occurrences of a measurement of Influenza test measurements excluding antibody testing       - value as concept is any of: Detected; Detected; Positive; Positive; Present; Present     - where event starts between 21 days Before and all days After index start date and event starts between all days Before and 0 days After index end date     - or at least 1 occurrences of an observation of Influenza test measurements excluding antibody testing       - value as concept is any of: Detected; Detected; Positive; Positive; Present; Present     - where event starts between 21 days Before and all days After index start date and event starts between all days Before and 0 days After index end date - where event starts between 180 days Before and 1 days Before index start date | <https://atlas.ohdsi.org/#/cohortdefinition/212> |
| **Comorbidities** |  |  |
| Asthma | People having any of the following:   - a drug exposure of Asthma therapy   - with age < 55 - Having all of the following criteria:   - at least 1 occurrences of a drug exposure of Asthma therapy   - where event starts between 365 days Before and 180 days Before index start date - a condition occurrence of Asthma - an observation of Asthma   with continuous observation of at least 0 days prior and 0 days after event index date; and limit initial events to: earliest event per person. | <https://atlas.ohdsi.org/#/cohortdefinition/218> |
| Autoimmune condition | People having any of the following condition occurrence/observation:   - Type 1 Diabetes Mellitus; Rheumatoid arthritis; Psoriasis; Psoriatic Arthritis; Multiple sclerosis; Systemic lupus erythematosus; Addison's disease; Graves' disease; Sjogren's syndrome; Hashimoto thyroiditis; Myasthenia gravis; Vasculitis; Pernicious anemia; Celiac disease; Scleroderma; Sarcoidosis; Ulcerative colitis; Crohn's disease   with continuous observation of at least 0 days prior and 0 days after event index date; and limit initial events to: earliest event per person. | <https://atlas.ohdsi.org/#/cohortdefinition/220> |
| Chronic kidney disease | People having any of the following:   - a condition occurrence of Chronic kidney disease   with continuous observation of at least 0 days prior and 0 days after event index date; and limit initial events to: earliest event per person.  For people matching the Primary Events; include:  Having any of the following criteria:   - at least 1 occurrences of a condition occurrence of Chronic kidney disease where event starts between 30 days After and all days After index start date - or at least 2 occurrences of a procedure of Dialysis - where event starts between 0 days Before and all days After index start date - or at least 2 occurrences of an observation of Dialysis - where event starts between 0 days Before and all days After index start date - or at least 2 occurrences of a condition occurrence of Dialysis - where event starts between 0 days Before and all days After index start date | <https://atlas.ohdsi.org/#/cohortdefinition/228> |
| Chronic obstructive pulmonary disease | People having any of the following:   - a drug exposure of COPD combo-therapy   - with age >= 55 - a drug exposure of COPD mono-therapy   - with age >= 55 - a condition occurrence of Chronic obstructive lung disease   with continuous observation of at least 0 days prior and 0 days after event index date; and limit initial events to: earliest event per person. | <https://atlas.ohdsi.org/#/cohortdefinition/219> |
| Dementia | People having any of the following:   - a condition occurrence of Dementia   with continuous observation of at least 0 days prior and 0 days after event index date; and limit initial events to: earliest event per person. | <https://atlas.ohdsi.org/#/cohortdefinition/226> |
| Heart disease | People having any of the following:   - a condition occurrence of Heart disease conditions   with continuous observation of at least 0 days prior and 0 days after event index date; and limit initial events to: earliest event per person. | <https://atlas.ohdsi.org/#/cohortdefinition/231> |
| Hypertension | People having any of the following:   - a condition occurrence of Hypertension   with continuous observation of at least 0 days prior and 0 days after event index date; and limit initial events to: earliest event per person. | <https://atlas.ohdsi.org/#/cohortdefinition/227> |
| History of cancer (malignant neoplasm excluding non-melanoma skin cancer) | People having any of the following:   - a condition occurrence of Malignant neoplasms excluding non-melanoma skin cancer - an observation of Malignant neoplasms excluding non-melanoma skin cancer   with continuous observation of at least 0 days prior and 0 days after event index date; and limit initial events to: earliest event per person. | <https://atlas.ohdsi.org/#/cohortdefinition/222> |
| Obesity | People having any of the following:   - a measurement of body mass index (BMI) measurement with value as number between 30 and 60 (inclusive) - a condition occurrence of obesity diagnoses - an observation of obesity diagnoses - a measurement of body weight   - with value as number between 120 and 200 (inclusive)   - unit is any of: kilogram - an observation of body weight   - with value as number between 120 and 200 (inclusive)   - unit is any of: kilogram   with continuous observation of at least 0 days prior and 0 days after event index date; and limit initial events to: earliest event per person. | <https://atlas.ohdsi.org/#/cohortdefinition/224> |
| Type 2 Diabetes Mellitus | People having any of the following:   - a condition occurrence of Type 2 Diabetes Mellitus - an observation of History of diabetes   with continuous observation of at least 0 days prior and 0 days after event index date; and limit initial events to: earliest event per person. | <https://atlas.ohdsi.org/#/cohortdefinition/311> |
| **30-day outcomes** |  |  |
| Intensive services requirement | People having any of the following:   - a procedure of Mechanical ventilation - a measurement of Mechanical ventilation - an observation of Mechanical ventilation - a procedure of tracheostomy - a procedure of Extracorporeal membrane oxygenation (ECMO) procedure   with continuous observation of at least 0 days prior and 0 days after event index date; and limit initial events to: all events per person.  For people matching the Primary Events; include:  Having all of the following criteria:   - at least 1 occurrences of a visit occurrence of Inpatient Visit - where event starts between all days Before and 0 days After index start date and event ends between 0 days Before and all days After index start date. | <https://atlas.ohdsi.org/#/cohortdefinition/316> |
| ARDS during hospitalization | People having any of the following:   - a condition occurrence of Acute respiratory distress syndrome (ARDS) or Acute Respiratory Failure   with continuous observation of at least 0 days prior and 0 days after event index date; and limit initial events to: all events per person.  For people matching the Primary Events; include:  Having all of the following criteria:   - at least 1 occurrences of a visit occurrence of Inpatient Visit - where event starts between all days Before and 0 days After index start date and event ends between 0 days Before and all days After index start date | <https://atlas.ohdsi.org/#/cohortdefinition/278> |
| Cardiovascular disease events | People having any of the following:   - a condition occurrence of Acute myocardial Infarction - a condition occurrence of Sudden cardiac death6 - a condition occurrence of Ischemic stroke - a condition occurrence of intracranial bleed Hemorrhagic stroke - a condition occurrence of Heart Failure   with continuous observation of at least 0 days prior and 0 days after event index date; and limit initial events to: all events per person.  For people matching the Primary Events; include:  Having all of the following criteria:   - at least 1 occurrences of a visit occurrence of Inpatient or Emergency room (ER) visit   where event starts between all days Before and 0 days After index start date and event ends between 0 days Before and all days After index start date | <https://atlas.ohdsi.org/#/cohortdefinition/246> |
| Deep vein thrombosis events | People having any of the following:   - a condition occurrence of non-Cerebral Deep Vein Thrombosis   with continuous observation of at least 0 days prior and 0 days after event index date; and limit initial events to: all events per person. | <https://atlas.ohdsi.org/#/cohortdefinition/273> |
| Pulmonary embolism events | People having any of the following:   - a condition occurrence of Pulmonary Embolism   with continuous observation of at least 0 days prior and 0 days after event index date; and limit initial events to: all events per person. | <https://atlas.ohdsi.org/#/cohortdefinition/274> |
| Acute kidney injury during hospitalization | People having any of the following:   - a condition occurrence of Acute kidney injury   with continuous observation of at least 0 days prior and 0 days after event index date; and limit initial events to: all events per person.  For people matching the Primary Events; include:  Having all of the following criteria:   - at least 1 occurrences of a visit occurrence of Inpatient Visit - where event starts between all days Before and 0 days After index start date and event ends between 0 days Before and all days After index start date | <https://atlas.ohdsi.org/#/cohortdefinition/276> |
| Sepsis during hospitalization | People having any of the following:   - a condition occurrence of Sepsis   with continuous observation of at least 0 days prior and 0 days after event index date; and limit initial events to: all events per person.  For people matching the Primary Events; include:  Having all of the following criteria:   - at least 1 occurrences of a visit occurrence of Inpatient Visit - where event starts between all days Before and 0 days After index start date and event ends between 0 days Before and all days After index start date | <https://atlas.ohdsi.org/#/cohortdefinition/277> |

Note: To access the webpage with all the definitions (<https://atlas.ohdsi.org>); users must sign in with an email address (all email addresses are accepted).

#### Appendix Table A3. SNOMED codes used to identify cancer types by anatomical location

| **Cancer** | **Condition code** | **Concept ID** |
| --- | --- | --- |
| Breast | Primary malignant neoplasm of Breast | 4162253 |
| Prostate | Primary malignant neoplasm of Prostate | 200962 |
| Colorectal | Malignant neoplasm of Colon and/or rectum | 36683531 |
| Bladder | Primary malignant neoplasm of Bladder | 196360 |
| Lung | Malignant tumor of Lung | 443388 |
| Melanoma | Malignant Melanoma | 4162276 |
| Thyroid gland | Primary malignant neoplasm of Thyroid gland | 133424 |
| Non-hodgkin's lymphoma | Non-hodgkin's lymphoma | 4038838 |
| Uterus | Primary malignant neoplasm of Uterus | 45770892 |
| Kidney | Primary malignant neoplasm of Kidney | 198985 |
| Leukemia | Leukemia | 317510 |
| Lip, oral cavity and pharynx | Malignant neoplasm of Lip, oral cavity and pharynx | 436045 |
| Central nervous system | Malignant neoplasm of Central nervous system | 4155285 |
| Ovary | Primary malignant neoplasm of ovary | 200051 |
| Multiple myeloma | Multiple myeloma | 437233 |
| Larynx | Primary malignant neoplasm of larynx | 26052 |
| Cervix | Primary malignant neoplasm of uterine cervix | 196359 |
| Stomach | Primary malignant neoplasm of stomach | 196044 |
| Hodgkin's disease | Hodgkin's disease | 4038835 |
| Testis | Primary malignant neoplasm of testis | 433716 |
| Pancreas | Primary malignant neoplasm of Pancreas | 199754 |
| Liver | Malignant neoplasm of Liver | 4246127 |
| Oropharynx | Primary malignant neoplasm of Oropharynx | 432833 |
| Vulva | Primary malignant neoplasm of vulva | 195197 |
| Esophagus | Primary malignant neoplasm of esophagus | 26638 |
| Gallbladder | Primary malignant neoplasm of gallbladder | 197806 |

Note: all descendants codes included can be consulted at: <https://athena.ohdsi.org/>

#### Appendix Table A4. Lifetime cancer prevalence, by study cohort and database

|  | **SIDIAP** | **CU-AMC-**  **HDC** | **CUIMC** | **HealthVerity** | **IQVIA-**  **OpenClaims** | **Optum-**  **EHR** | **STARR-**  **OMOP** | **VA-**  **OMOP** | **Total** |
| --- | --- | --- | --- | --- | --- | --- | --- | --- | --- |
| **Diagnosed with COVID-19** |  |  |  |  |  |  |  |  |  |
| All, n | 122,141 | 7,270 | 8,519 | 114,173 | 466,191 | 129,512 | 3,328 | 55,557 | 906,691 |
| With a history of cancer, n | 8,854 | 806 | 1,433 | 4,857 | 72,994 | 17,630 | 821 | 10,760 | 118,155 |
| Lifetime cancer prevalence, % (95%CI) | 7.2  (7.1-7.3) | 11.1  (10.4-11.8) | 16.8  (16-17.6) | 4.3  (4.2-4.4) | 15.7  (15.6-15.8) | 13.6  (13.4-13.8) | 24.7  (23.2-26.2) | 19.4  (19.1-19.7) | 13.0  (12.9-13.1) |
| **Hospitalized with COVID-19** |  |  |  |  |  |  |  |  |  |
| All, n | 18,202 | 1,434 | 2,600 | 7,581 | 133,091 | 22,024 | 615 | 10,471 | 196,018 |
| With a history of cancer, n | 2,610 | 265 | 561 | 797 | 29,628 | 4,451 | 244 | 3,383 | 41,939 |
| Lifetime cancer prevalence, % (95%CI) | 14.3  (13.8-14.8) | 18.5  (16.5-20.5) | 21.6  (20-23.2) | 10.5  (9.8-11.2) | 22.3  (22.1-22.5) | 20.2 (19.7-20.7) | 39.7  (35.8-43.6) | 32.3  (31.4-33.2) | 21.4  (21.2-21.6) |
| **Hospitalized with seasonal influenza in 2017-2018** |  |  |  |  |  |  |  |  |  |
| All, n | - | 853 | 655 | - | 229,544 | 2,213 | - | 9,441 | 242,706 |
| With a history of cancer, n | - | 219 | 179 | - | 57,529 | 473 | - | 3,677 | 62,077 |
| Lifetime cancer prevalence, % (95%CI) | - | 25.7  (22.8-28.6) | 27.3  (23.9-30.7) | - | 25.1  (24.9-25.3) | 21.4  (19.7-23.1) | - | 38.9  (37.9-39.9) | 25.6  (25.4-25.8) |

Abbreviations: COVID-19: Coronavirus Disease 2019; SIDIAP: Information System for Research in Primary Care; CU-AMC-HDC: Colorado University Anschutz Medical Campus Health Data Compass; CUIMC: Columbia University Irving Medical Center; STARR-OMOP: Stanford Medicine Research Data Repository; VA-OMOP:Department of Veterans Affairs.

#### Appendix Table A5. Proportion of demographics, comorbidities and outcomes among patients with a history of cancer diagnosed and hospitalized with COVID-19

| **Patients with a history of cancer diagnosed with COVID-19** | | | | | | | | |
| --- | --- | --- | --- | --- | --- | --- | --- | --- |
| **Characteristics, % (95%CI)** | **SIDIAP**  **n=8,854** | **CU-AMC-HDC**  **n=806** | **CUIMC**  **n=1,433** | **HealthVerity**  **n=4,857** | **IQVIA-**  **OpenClaims**  **n=72,994** | **Optum-EHR**  **n=17,630** | **STARR-OMOP**  **n=821** | **VA-OMOP**  **n=10,760** |
| Male | 46.1  (45.1-47.1) | 47.0  (43.6-50.4) | 45.3  (42.7-47.9) | 46.3  (44.9-47.7) | 46.8  (46.4-47.2) | 41.0  (40.3-41.7) | 53.0  (49.6-56.4) | 92.9  (92.4-93.4) |
| **Antineoplastic agents** |  |  |  |  |  |  |  |  |
| Year prior | 13.6  (12.9-14.3) | 35.1  (31.8-38.4) | 24.6  (22.4-26.8) | 20.3  (19.2-21.4) | 17.8  (17.5-18.1) | 16.5  (16.0-17.0) | 31.9  (28.7-35.1) | 21.5  (20.7-22.3) |
| Month prior | 10.9  (10.3-11.5) | 22.5  (19.6-25.4) | 12.3  (10.6-14.0) | 5.4  (4.8-6.0) | 9.5  (9.3-9.7) | 6.1  (5.7-6.5) | 18.8  (16.1-21.5) | 12.0  (11.4-12.6) |
| **Comorbidities** |  |  |  |  |  |  |  |  |
| Asthma | 4.7  (4.3-5.1) | 15.4  (12.9-17.9) | 20.5  (18.4-22.6) | 9.9  (9.1-10.7) | 18.5  (18.2-18.8) | 20.3  (19.7-20.9) | 14.3  (11.9-16.7) | 10.8  (10.2-11.4) |
| COPD | 33.7  (32.7-34.7) | 23.7  (20.8-26.6) | 19.8  (17.7-21.9) | 17.3  (16.2-18.4) | 28.1  (27.8-28.4) | 18.1  (17.5-18.7) | 11.9  (9.7-14.1) | 43.3  (42.4-44.2) |
| Type 2 diabetes | 17.8  (17.0-18.6) | 26.9  (23.8-30.0) | 38.0  (35.5-40.5) | 31.7  (30.4-33.0) | 52.2  (51.8-52.6) | 30.2  (29.5-30.9) | 22.0  (19.2-24.8) | 50.3  (49.4-51.2) |
| Hyperlipidemia | 22.8  (21.9-23.7) | 38.7  (35.3-42.1) | 30.8  (28.4-33.2) | 40.1  (38.7-41.5) | 40.0  (39.6-40.4) | 44.4  (43.7-45.1) | 38.7  (35.4-42.0) | 58.8  (57.9-59.7) |
| Obesity | 41.8  (40.8-42.8) | 49.3  (45.8-52.8) | 52.8  (50.2-55.4) | 21.8  (20.6-23.0) | 36.0  (35.7-36.3) | 59.6  (58.9-60.3) | 40.0  (36.6-43.4) | 52.7  (51.8-53.6) |
| Heart disease | 34.3  (33.3-35.3) | 50.0  (46.5-53.5) | 62.8  (60.3-65.3) | 39.9  (38.5-41.3) | 72.7  (72.4-73.0) | 51.1  (50.4-51.8) | 40.8  (37.4-44.2) | 71.4  (70.5-72.3) |
| Hypertension | 33.2  (32.2-34.2) | 60.2  (56.8-63.6) | 70.0  (67.6-72.4) | 58.1  (56.7-59.5) | 83.6  (83.3-83.9) | 61.2  (60.5-61.9) | 52.4  (49.0-55.8) | 86.7  (86.1-87.3) |
| Anxiety | 24.2  (23.3-25.1) | 19.1  (16.4-21.8) | 12.2  (10.5-13.9) | 13.5  (12.5-14.5) | 13.8  (13.5-14.1) | 18.0  (17.4-18.6) | 16.6  (14.1-19.1) | 31.1  (30.2-32.0) |
| Dementia | 10.7  (10.1-11.3) | 5.8  (4.2-7.4) | 11.0  (9.4-12.6) | 7.1  (6.4-7.8) | 21.4  (21.1-21.7) | 6.8  (6.4-7.2) | 1.7  (0.8-2.6) | 14.3  (13.6-15.0) |
| Depression | 7.2  (6.7-7.7) | 9.2  (7.2-11.2) | 14.8  (13.0-16.6) | 2.7  (2.2-3.2) | 12.0  (11.8-12.2) | 10.5  (10.0-11.0) | 10.4  (8.3-12.5) | 21.1  (20.3-21.9) |
| Anemia | 19.6  (18.8-20.4) | 22.1  (19.2-25.0) | 19.3  (17.3-21.3) | 24.8  (23.6-26.0) | 29.0  (28.7-29.3) | 18.8  (18.2-19.4) | 24.7  (21.7-27.7) | 25.8  (25.0-26.6) |
| Autoimmune condition | 10.4  (9.8-11.0) | 19.9  (17.1-22.7) | 30.8  (28.4-33.2) | 13.0  (12.1-13.9) | 35.8  (35.5-36.1) | 18.4  (17.8-19.0) | 14.6  (12.2-17.0) | 30.3  (29.4-31.2) |
| Chronic kidney disease | 19.4  (18.6-20.2) | 23.8  (20.9-26.7) | 27.4  (25.1-29.7) | 18.4  (17.3-19.5) | 34.1  (33.8-34.4) | 29.3  (28.6-30.0) | 18.0  (15.4-20.6) | 34.1  (33.2-35.0) |
| Chronic liver disease | 1.9  (1.6-2.2) | 3.2  (2.0-4.4) | 3.6  (2.6-4.6) | 2.2  (1.8-2.6) | 2.2  (2.1-2.3) | 2.5  (2.3-2.7) | 7.3  (5.5-9.1) | 6.1  (5.6-6.6) |
| **Outcomes** |  |  |  |  |  |  |  |  |
| Death | 14.4  (13.7-15.1) | - | 10.4  (8.8-12.0) | - | - | 1.9  (1.7-2.1) | - | 7.6  (7.1-8.1) |
| Hospitalization | 24.9  (24.0-25.8) | - | 34.8  (32.3-37.3) | 13.5  (12.5-14.5) | 39.3  (38.9-39.7) | 24.1  (23.5-24.7) | 27.0  (24.0-30.0) | 27.1  (26.3-27.9) |
| **Patients with a history of cancer hospitalized with COVID-19** | | | | | | | | |
| **Characteristics, % (95%CI)** | **SIDIAP**  **n=2,610** | **CU-AMC-HDC**  **n=265** | **CUIMC**  **n=561** | **HealthVerity**  **n=797** | **IQVIA-**  **OpenClaims**  **n=29,628** | **Optum-EHR**  **n=4,451** | **STARR-OMOP**  **n=244** | **VA-OMOP**  **n=3,383** |
| Male | 60.3  (58.4-62.2) | 53.2  (47.2-59.2) | 53.7  (49.6-57.8) | 49.9  (46.4-53.4) | 53.2  (52.6-53.8) | 49.5  (48.0-51.0) | 58.2  (52.0-64.4) | 96.4  (95.8-97.0) |
| **Antineoplastic agents,** |  |  |  |  |  |  |  |  |
| Year prior | 13.7  (12.4-15.0) | 37.7  (31.8-43.6) | 26.0  (22.4-29.6) | 18.6  (15.9-21.3) | 19.8  (19.3-20.3) | 21.7  (20.5-22.9) | 29.9  (24.1-35.7) | 24.1  (22.7-25.5) |
| Month prior | 10.8  (9.6-12.0) | 28.3  (22.9-33.7) | 14.6  (11.7-17.5) | 5.9  (4.3-7.5) | 10.6  (10.2-11.0) | 11.3  (10.4-12.2) | 18.9  (14.0-23.8) | 14.2  (13.0-15.4) |
| **Comorbidities** |  |  |  |  |  |  |  |  |
| Asthma | 3.9  (3.2-4.6) | 12.1  (8.2-16.0) | 22.8  (19.3-26.3) | 10.0  (7.9-12.1) | 17.0  (16.6-17.4) | 16.4  (15.3-17.5) | 16.4  (11.7-21.1) | 9.9  (8.9-10.9) |
| COPD | 41.5  (39.6-43.4) | 29.4  (23.9-34.9) | 28.2  (24.5-31.9) | 31.4  (28.2-34.6) | 33.7  (33.2-34.2) | 28.0  (26.7-29.3) | 11.1  (7.2-15.0) | 53.2  (51.5-54.9) |
| Type 2 diabetes | 22.5  (20.9-24.1) | 36.2  (30.4-42.0) | 55.1  (51.0-59.2) | 43.0  (39.6-46.4) | 59.4  (58.8-60.0) | 40.7  (39.3-42.1) | 23.8  (18.4-29.2) | 58.7  (57.0-60.4) |
| Hyperlipidemia | 24.6  (22.9-26.3) | 42.3  (36.3-48.3) | 38.0  (34.0-42.0) | 48.4  (44.9-51.9) | 43.1  (42.5-43.7) | 48.7  (47.2-50.2) | 44.3  (38.1-50.5) | 61.2  (59.6-62.8) |
| Obesity | 47.8  (45.9-49.7) | 52.1  (46.1-58.1) | 57.0  (52.9-61.1) | 25.7  (22.7-28.7) | 38.5  (37.9-39.1) | 59.9  (58.5-61.3) | 39.8  (33.7-45.9) | 53.4  (51.7-55.1) |
| Heart disease | 41.6  (39.7-43.5) | 59.2  (53.3-65.1) | 76.5  (73.0-80.0) | 60.9  (57.5-64.3) | 79.6  (79.1-80.1) | 63.4  (62.0-64.8) | 45.5  (39.2-51.8) | 79.1  (77.7-80.5) |
| Hypertension | 37.3  (35.4-39.2) | 69.4  (63.8-75.0) | 83.1  (80.0-86.2) | 74.4  (71.4-77.4) | 89.5  (89.2-89.8) | 73.2  (71.9-74.5) | 58.2  (52.0-64.4) | 92.8  (91.9-93.7) |
| Anxiety | 19.3  (17.8-20.8) | 18.1  (13.5-22.7) | 10.7  (8.1-13.3) | 19.4  (16.7-22.1) | 13.3  (12.9-13.7) | 17.5  (16.4-18.6) | 18.9  (14.0-23.8) | 30.1  (28.6-31.6) |
| Dementia | 7.2  (6.2-8.2) | 7.5  (4.3-10.7) | 21.2  (17.8-24.6) | 15.8  (13.3-18.3) | 21.8  (21.3-22.3) | 11.1  (10.2-12.0) | - | 23.7  (22.3-25.1) |
| Depression | 5.7  (4.8-6.6) | 9.1  (5.6-12.6) | 20.0  (16.7-23.3) | 4.9  (3.4-6.4) | 11.6  (11.2-12.0) | 9.2  (8.4-10.0) | 12.7  (8.5-16.9) | 19.1  (17.8-20.4) |
| Anemia | 20.0  (18.5-21.5) | 35.1  (29.3-40.9) | 25.0  (21.4-28.6) | 40.9  (37.5-44.3) | 35.5  (35.0-36.0) | 30.0  (28.7-31.3) | 32.0  (26.1-37.9) | 36.7  (35.1-38.3) |
| Autoimmune condition | 11.2  (10.0-12.4) | 20.0  (15.2-24.8) | 38.1  (34.1-42.1) | 17.8  (15.1-20.5) | 39.0  (38.4-39.6) | 20.6  (19.4-21.8) | 11.9  (7.8-16.0) | 35.4  (33.8-37.0) |
| Chronic kidney disease | 23.6  (22.0-25.2) | 34.0  (28.3-39.7) | 40.8  (36.7-44.9) | 36.0  (32.7-39.3) | 43.8  (43.2-44.4) | 40.8  (39.4-42.2) | 20.1  (15.1-25.1) | 44.3  (42.6-46.0) |
| Chronic liver disease | 1.8  (1.3-2.3) | 4.9  (2.3-7.5) | 5.0  (3.2-6.8) | 4.0  (2.6-5.4) | 3.0  (2.8-3.2) | 3.7  (3.1-4.3) | 4.9  (2.2-7.6) | 8.8  (7.8-9.8) |
| **Outcomes** |  |  |  |  |  |  |  |  |
| Death | 21.4  (19.8-23.0) | - | 26.2  (22.6-29.8) | - | - | 6.2  (5.5-6.9) | - | 18.1  (16.8-19.4) |
| Intensive services | - | - | - | 6.3  (4.6-8.0) | 9.7  (9.4-10.0) | 13.6  (12.6-14.6) | 5.7  (2.8-8.6) | 16.0  (14.8-17.2) |
| ARDS | - | - | 15.9  (12.9-18.9) | 26.5  (23.4-29.6) | 31.2  (30.7-31.7) | 40.2  (38.8-41.6) | 8.2  (4.7-11.7) | 41.2  (39.5-42.9) |
| Cardiovascular events | 1.1  (0.7-1.5) | - | 6.8  (4.7-8.9) | 10.8  (8.6-13.0) | 8.7  (8.4-9.0) | 16.7  (15.6-17.8) | 8.2  (4.7-11.7) | 20.8  (19.4-22.2) |
| Deep vein thrombosis | - | - | 2.1  (0.9-3.3) | 3.0  (1.8-4.2) | 2.1  (1.9-2.3) | 4.4  (3.8-5.0) | - | 4.8  (4.1-5.5) |
| Pulmonary embolism | 0.7  (0.4-1.0) | - | 2.7  (1.4-4.0) | 2.1  (1.1-3.1) | 1.6  (1.5-1.7) | 3.6  (3.1-4.1) | - | 4.0  (3.3-4.7) |
| Acute kidney injury | 0.2  (0.0-0.4) | - | 16.0  (13.0-19.0) | 11.4  (9.2-13.6) | 10.2  (9.9-10.5) | 16.4  (15.3-17.5) | 11.9  (7.8-16.0) | 14.4  (13.2-15.6) |
| Sepsis | - | - | 6.1  (4.1-8.1) | 17.7  (15.0-20.4) | 17.1  (16.7-17.5) | 24.5  (23.2-25.8) | 9.8  (6.1-13.5) | 20.5  (19.1-21.9) |

Note: - means data not available or below the minimum cell count required (5 individuals)

Abbreviations: COVID-19: Coronavirus disease 2019; COPD: Chronic Obstructive Pulmonary Disease; ARDS: Acute Respiratory Distress Syndrome; SIDIAP: Information System for Research in Primary Care; CU-AMC-HDC: Colorado University Anschutz Medical Campus Health Data Compass; CUIMC: Columbia University Irving Medical Center; STARR-OMOP: Stanford Medicine Research Data Repository; VA-OMOP:Department of Veterans Affairs

#### Appendix Figure A2. Age distribution among patients with a history of cancer diagnosed and hospitalized with COVID-19

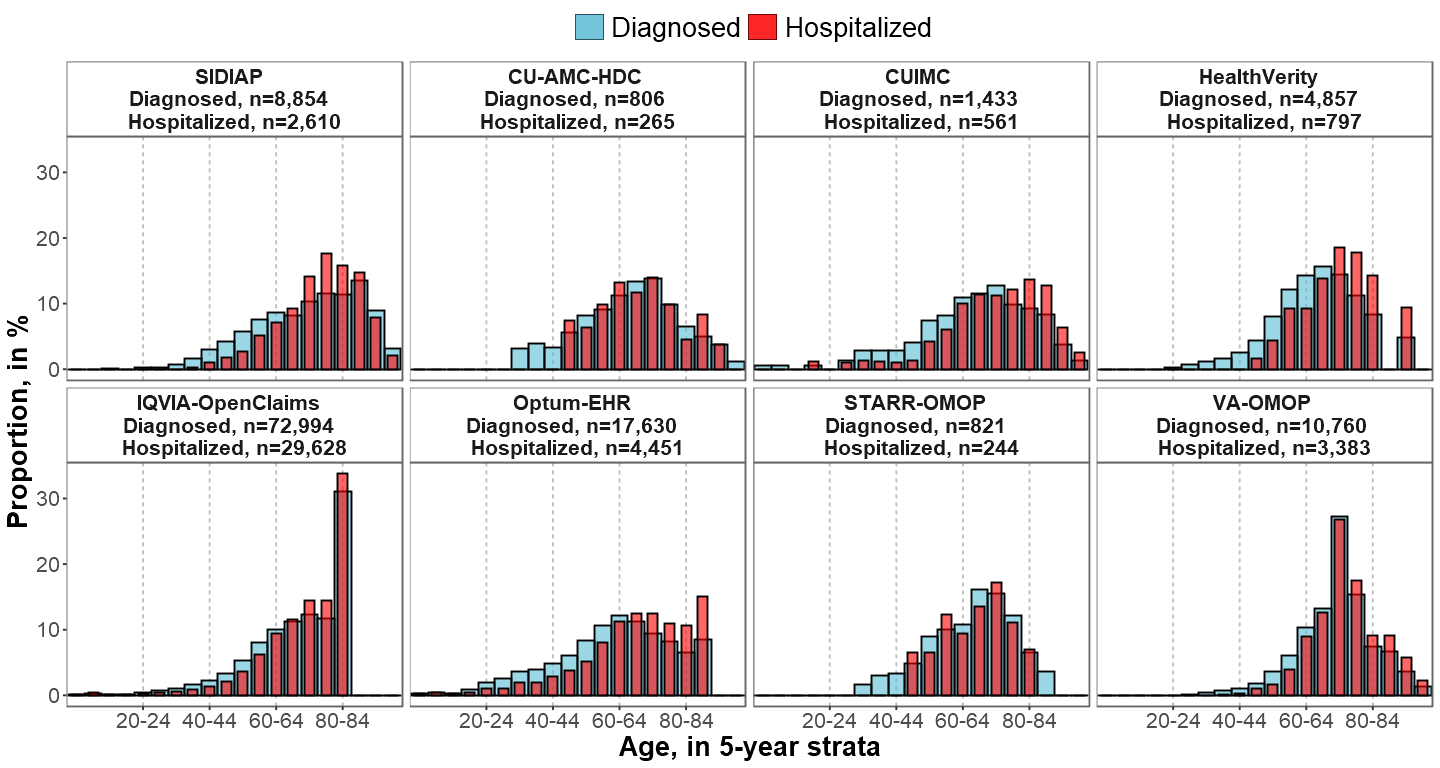

Note: In IQVIA-OpenClaims; the age category 80-84 equals to “80 years or older”

Abbreviations: COVID-19: Coronavirus disease 2019; SIDIAP: Information System for Research in Primary Care; CU-AMC-HDC: Colorado University Anschutz Medical Campus Health Data Compass; CUIMC: Columbia University Irving Medical Center; STARR-OMOP: Stanford Medicine Research Data Repository; VA-OMOP:Department of Veterans Affairs

#### Appendix Table A6. Proportion and rank of cancer types among patients with a history of cancer *diagnosed* and *hospitalized* with COVID-19

Cancer types ordered in descending order according to the most 5-year prevalent cancer types in the US based on data from the International Agency for Cancer Research.^16^

| **Patients with a history of cancer *diagnosed* with COVID-19** | | | | | | | | | | | | | | | | |
| --- | --- | --- | --- | --- | --- | --- | --- | --- | --- | --- | --- | --- | --- | --- | --- | --- |
|  | **SIDIAP**  **n=8,854** | | **CU-AMC-HDC**  **n=806** | | **CUIMC**  **n=1,433** | | **HealthVerity**  **n=4,857** | | **IQVIA-**  **OpenClaims**  **n=72,994** | | **Optum-EHR**  **n=17,630** | | **STARR-OMOP**  **n=821** | | **VA-OMOP**  **n=10,760** | |
| **Cancer type** | **%** | **Top 10 rank** | **%** | **Top 10 rank** | **%** | **Top 10 rank** | **%** | **Top 10 rank** | **%** | **Top 10 rank** | **%** | **Top 10 rank** | **%** | **Top 10 rank** | **%** | **Top 10 rank** |
| Breast | 14.2 | 1 | 7.3 | 1 | 5.2 | 3 | 11.5 | 2 | 6.0 | 2 | 6.7 | 1 | 12.3 | 1 | 0.8 |  |
| Prostate | 9.4 | 3 | 6.8 | 2 | 6.1 | 1 | 12.2 | 1 | 7.1 | 1 | 4.9 | 2 | 10.2 | 2 | 18.1 | 1 |
| Colorectal | 10.4 | 2 | 2.9 | 7 | 2.7 | 8 | 3.9 | 4 | 2.6 | 6 | 1.8 | 8 | 3.7 | 9 | 2.6 | 7 |
| Bladder | 6.4 | 4 | 2.2 | 9 | 1.7 |  | 2.5 | 8 | 1.7 | 7 | 1.4 | 10 | 3.2 | 10 | 3.3 | 4 |
| Lung | 0.6 |  | 3.8 | 4 | 3.1 | 6 | 2.7 | 7 | 2.7 | 4 | 2.2 | 7 | 6.9 | 4 | 3.9 | 2 |
| Melanoma | 2.9 | 6 | 3.0 | 6 | 0.6 |  | 1.0 |  | 0.6 |  | 1.1 |  | 2.1 |  | 1.1 |  |
| Thyroid | 1.4 |  | 2.2 | 10 | 1.7 |  | 3.5 | 5 | 1.1 |  | 1.5 | 9 | 5.2 | 6 | 0.7 |  |
| NHL | 3.0 | 5 | 5.0 | 3 | 5.4 | 2 | 4.6 | 3 | 3.5 | 3 | 2.6 | 5 | 5.8 | 5 | 3.8 | 3 |
| Uterus | 2.5 | 8 | 1.6 |  | 2.0 | 9 | 1.9 |  | 1.1 |  | 3.7 | 3 | 2.2 |  | 0.1 |  |
| Kidney | 2.4 | 9 | 1.4 |  | 1.8 | 10 | 2.1 | 10 | 1.4 | 10 | 1.2 |  | 3.0 |  | 2.5 | 8 |
| Leukemia | 2.7 | 7 | 3.3 | 5 | 4.4 | 4 | 2.8 | 6 | 2.6 | 5 | 2.3 | 6 | 3.8 | 8 | 2.6 | 6 |
| Lip, oral cavity and pharynx | 1.4 | 10 | 1.7 |  | 1.0 |  | 1.8 |  | 1.2 |  | 3.7 | 4 | 4.9 | 7 | 2.9 | 5 |
| Central nervous system | 0.9 |  | - |  | 1.7 |  | 1.4 |  | 1.2 |  | 0.8 |  | 2.4 |  | 0.8 |  |
| Ovary | 0.8 |  | - |  | 0.7 |  | 0.9 |  | 0.5 |  | 0.5 |  | 1.2 |  | - |  |
| Multiple myeloma | 0.9 |  | 2.5 | 8 | 3.1 | 7 | 2.1 | 9 | 1.6 | 8 | 0.9 |  | 1.8 |  | 1.2 |  |
| Larynx | 0.7 |  | - |  | - |  | 0.6 |  | 0.5 |  | 0.3 |  | 1.6 |  | 1.3 | 10 |
| Cervix | 0.7 |  | - |  | 0.6 |  | 0.5 |  | 0.3 |  | 0.7 |  | - |  | - |  |
| Oropharynx | 0.1 |  | - |  | - |  | 0.5 |  | 0.4 |  | 0.3 |  | 1.6 |  | 1.2 |  |
| Stomach | 1.0 |  | - |  | - |  | 0.7 |  | 0.4 |  | 0.3 |  | 1.2 |  | 0.4 |  |
| Hodgkin's disease | 0.5 |  | - |  | 0.6 |  | 0.8 |  | 0.5 |  | 0.5 |  | 1.3 |  | 0.6 |  |
| Testis | 0.4 |  | - |  | - |  | 0.4 |  | 0.1 |  | 1.2 |  | - |  | 0.3 |  |
| Pancreas | 0.8 |  | 1.2 |  | 1.2 |  | 0.9 |  | 0.7 |  | 0.4 |  | 2.9 |  | 0.6 |  |
| Liver | 1.2 |  | 2.0 |  | 3.3 | 5 | 1.3 |  | 1.5 | 9 | 1.3 |  | 7.7 | 3 | 1.8 | 9 |
| **Patients with a history of cancer *hospitalized* with COVID-19** | | | | | | | | | | | | | | | |  |
|  | **SIDIAP**  **n=2,610** | | **CU-AMC-HDC**  **n=265** | | **CUIMC**  **n=561** | | **HealthVerity**  **n=797** | | **IQVIA-**  **OpenClaims**  **n=29,628** | | **Optum-EHR**  **n=4,451** | | **STARR-OMOP**  **n=244** | | **VA-OMOP**  **n=3,383** | |
| **Cancer type** | **%** | **Top 10 rank** | **%** | **Top 10 rank** | **%** | **Top 10 rank** | **%** | **Top 10 rank** | **%** | **Top 10 rank** | **%** | **Top 10 rank** | **%** | **Top 10 rank** | **%** | **Top 10 rank** |
| Breast | 9.0 | 3 | 3.8 | 6 | 3.2 | 8 | 9.2 | 2 | 5.4 | 2 | 6.4 | 1 | 7.4 | 4 | 0.6 |  |
| Prostate | 12.8 | 1 | 6.4 | 1 & 2 | 7.7 | 1 | 10.8 | 1 | 8.5 | 1 | 6.0 | 2 | 10.7 | 1 | 19.4 | 1 |
| Colorectal | 11.9 | 2 | - |  | 3.4 | 6 | 5.0 | 4 | 3.0 | 6 | 2.5 | 9 | - |  | 3.2 | 6 |
| Bladder | 8.5 | 4 | - |  | 1.1 |  | 3.0 | 8 | 2.0 | 9 | 2.4 | 10 | - |  | 3.9 | 4 |
| Lung | 0.9 |  | 6.0 | 3 | 3.7 | 4 | 4.0 | 6 | 4.0 | 4 | 3.6 | 4 | 9.0 | 3 | 5.7 | 2 |
| Melanoma | 2.3 | 8 | - |  | - |  | 1.1 |  | 0.7 |  | 1.1 |  | - |  | 0.9 |  |
| Thyroid | 0.9 |  | - |  | - |  | 1.5 |  | 0.7 |  | 1.1 |  | 6.1 | 6 | 0.4 |  |
| NHL | 4.3 | 5 | 6.4 | 1 & 2 | 6.4 | 2 | 7.4 | 3 | 4.5 | 3 | 4.2 | 3 | - |  | 4.6 | 3 |
| Uterus | 1.8 | 9 | - |  | 1.8 | 10 | 2.8 | 9 | 1.1 |  | 3.0 | 7 | - |  | - |  |
| Kidney | 2.8 | 7 | - |  | 1.4 |  | 2.1 |  | 1.7 | 10 | 1.8 |  | - |  | 2.7 | 9 |
| Leukemia | 4.2 | 6 | 4.2 | 5 | 4.1 | 3 | 4.1 | 5 | 3.5 | 5 | 3.4 | 6 | 4.9 | 8 | 3.3 | 5 |
| Lip, oral cavity and pharynx | 1.4 |  | - |  | - |  | 2.5 | 10 | 1.4 |  | 3.5 | 5 | 9.0 | 2 | 2.8 | 8 |
| Central nervous system | 0.7 |  | - |  | 2.0 | 9 | 1.5 |  | 1.6 |  | 1.6 |  | - |  | 1.3 |  |
| Ovary | 0.5 |  | - |  | - |  | - |  | 0.5 |  | 0.5 |  | - |  | - |  |
| Multiple myeloma | 1.2 |  | 4.2 | 4 | 3.4 | 7 | 3.4 | 7 | 2.2 | 8 | 1.7 |  | - |  | 1.8 | 10 |
| Larynx | 1.2 |  | - |  | - |  | 1.0 |  | 0.7 |  | 0.4 |  | - |  | 1.4 |  |
| Cervix | 0.4 |  | - |  | - |  | 0.8 |  | 0.3 |  | 0.7 |  | - |  | - |  |
| Oropharynx |  |  | - |  | - |  | - |  | 0.4 |  | 0.4 |  | 4.5 | 9 | 1.0 |  |
| Stomach | 0.9 |  | - |  | - |  | - |  | 0.5 |  | 0.5 |  | - |  | 0.4 |  |
| Hodgkin's disease | 0.2 |  | - |  | - |  | 1.5 |  | 0.6 |  | 0.7 |  | - |  | 0.6 |  |
| Testis | - |  | - |  | - |  | - |  | 0.1 |  | 1.0 |  | - |  | - |  |
| Pancreas | 0.9 |  | - |  | 1.2 |  | 0.9 |  | 0.7 |  | 0.7 |  | 5.3 | 7 | 0.8 |  |
| Liver | 1.5 | 10 | 3.8 | 7 | 3.6 | 5 | 2.3 |  | 2.2 | 7 | 2.7 | 8 | 6.6 | 5 | 2.8 | 7 |

Notes:The 10 most frequent cancer types are ranked in descending order by cohort and database from 1 to 10. In bold; hematological malignancies.

Abbreviations: COVID-19: Coronavirus disease 2019; NHL: Non-Hodgkin's lymphoma ; SIDIAP: Information System for Research in Primary Care; CU-AMC-HDC: Colorado University Anschutz Medical Campus Health Data Compass; CUIMC: Columbia University Irving Medical Center; STARR-OMOP: Stanford Medicine Research Data Repository; VA-OMOP:Department of Veterans Affairs.

#### Appendix Figure A3. Standardized mean differences of cancer types between patients with a history of cancer *diagnosed* and *hospitalized* with COVID-19.

SMD<0 means prevalence was greater in patients *diagnosed*; SMD>0 means prevalence was greater in patients *hospitalized.*

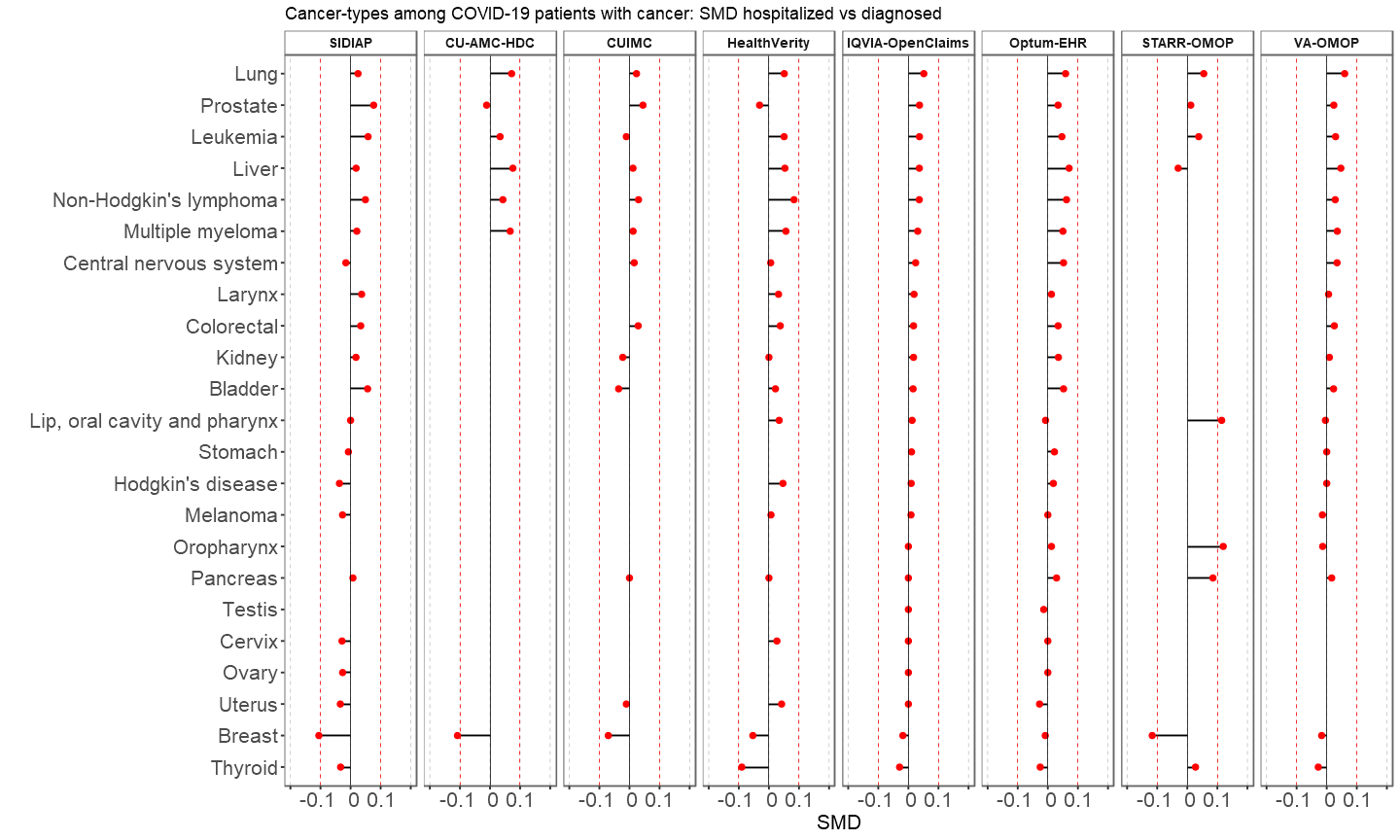

Notes: SMD calculated for cancer types meeting the minimum count required (5 individuals) in each database and cohort. Cancer types ordered according to SMD descending values in IQVIA-OpenClaims. Abbreviations: COVID-19: Coronavirus Disease 2019; SIDIAP: Information System for Research in Primary Care; CU-AMC-HDC: Colorado University Anschutz Medical Campus Health Data Compass; CUIMC: Columbia University Irving Medical Center; STARR-OMOP: Stanford Medicine Research Data Repository; VA-OMOP:Department of Veterans Affairs; SMD: Standardized mean difference

#### Appendix Table A7. Top 10 cancer types among patients with a history of cancer hospitalized with seasonal influenza (2017-2018)

| **Rank** | **CU-AMC-HDC**  **n= 219** | | **CUIMC**  **n=179** | | **IQVIA-OpenClaims**  **n= 57,529** | | **Optum-EHR**  **n= 473** | | **VA-OMOP**  **n= 3,677** | |
| --- | --- | --- | --- | --- | --- | --- | --- | --- | --- | --- |
|  | **cancer type** | **% (95%CI)** | **cancer type** | **% (95%CI)** | **cancer type** | **% (95%CI)** | **cancer type** | **% (95%CI)** | **cancer type** | **% (95%CI)** |
| 1 | Breast | 9.1  (5.3-12.9) | Leukemia | 10.6  (6.1-15.1) | Prostate | 7.1  (6.9-7.3) | Lip, oral cavity and pharynx | 10.1  (7.4-12.8) | Prostate | 12.9  (11.8-14.0) |
| 2 | Prostate | 7.8  (4.3-11.3) | NHL | 7.3  (3.5-11.1) | Breast | 6.4  (6.2-6.6) | Breast | 6.3  (4.1-8.5) | Lung | 6.2  (5.4-7.0) |
| 3 | Lung | 6.8  (3.4-10.2) | Colorrectal | 7.3  (3.5-11.1) | NHL | 5.6  (5.4-5.8) | Prostate | 4.7  (2.8-6.6) | NHL | 5.2  (4.5-5.9) |
| 4 | Leukemia | 6.4  (3.2-9.6) | Liver | 6.7  (3.0-10.4) | Lung | 5.1  (4.9-5.3) | Uterus | 4.7  (2.8-6.6) | Leukemia | 3.6  (3.0-4.2) |
| 5 | NHL | 6.4  (3.2-9.6) | Multiple myeloma | 6.7  (3.0-10.4) | Leukemia | 4.2  (4.0-4.4) | Lung | 4.4  (2.5-6.3) | Bladder | 3.6  (3.0-4.2) |
| 6 | Liver | 5.0  (2.1-7.9) | Lung | 5.6  (2.2-9.0) | Multiple myeloma | 3.1  (3.0-3.2) | Leukemia | 4.0  (2.2-5.8) | Lip, oral cavity and pharynx | 3.2  (2.6-3.8) |
| 7 |  |  | Prostate | 4.5  (1.5-7.5) | Colorrectal | 3.1  (3.0-3.2) | NHL | 3.6  (1.9-5.3) | Multiple myeloma | 2.7  (2.2-3.2) |
| 8 |  |  | Kidney | 3.9  (1.1-6.7) | Lip, oral cavity and pharynx | 2.4  (2.3-2.5) | Kidney | 2.1  (0.8-3.4) | Colorrectal | 2.4  (1.9-2.9) |
| 9 |  |  | Central nervous system | 3.9  (1.1-6.7) | Bladder | 2.3  (2.2-2.4) | Colorrectal | 1.9  (0.7-3.1) | Kidney | 20. (1.6-2.6) |
| 10 |  |  | Breast | 3.4  (0.8-6.0) | Liver | 2.0  (1.9-2.1) | Multiple myeloma | 1.7  (0.5-2.9) | Liver | 2.1  (1.6-2.6) |

Abbreviations: CU-AMC-HDC: Colorado University Anschutz Medical Campus Health Data Compass; CUIMC: Columbia University Irving Medical Center; STARR-OMOP: Stanford Medicine Research Data Repository; VA-OMOP:Department of Veterans Affairs; NHL: Non-Hodgkin's Lymphoma

#### Appendix Figure A4. Standardized mean differences of cancer tybes between patients with a history of cancer hospitalized with COVID-19 and with seasonal influenza (2017-2018).

SMD<0 means prevalence of cancer was greater in patients with seasonal influenza, SMD>0 means prevalence was greater in patients with COVID-19

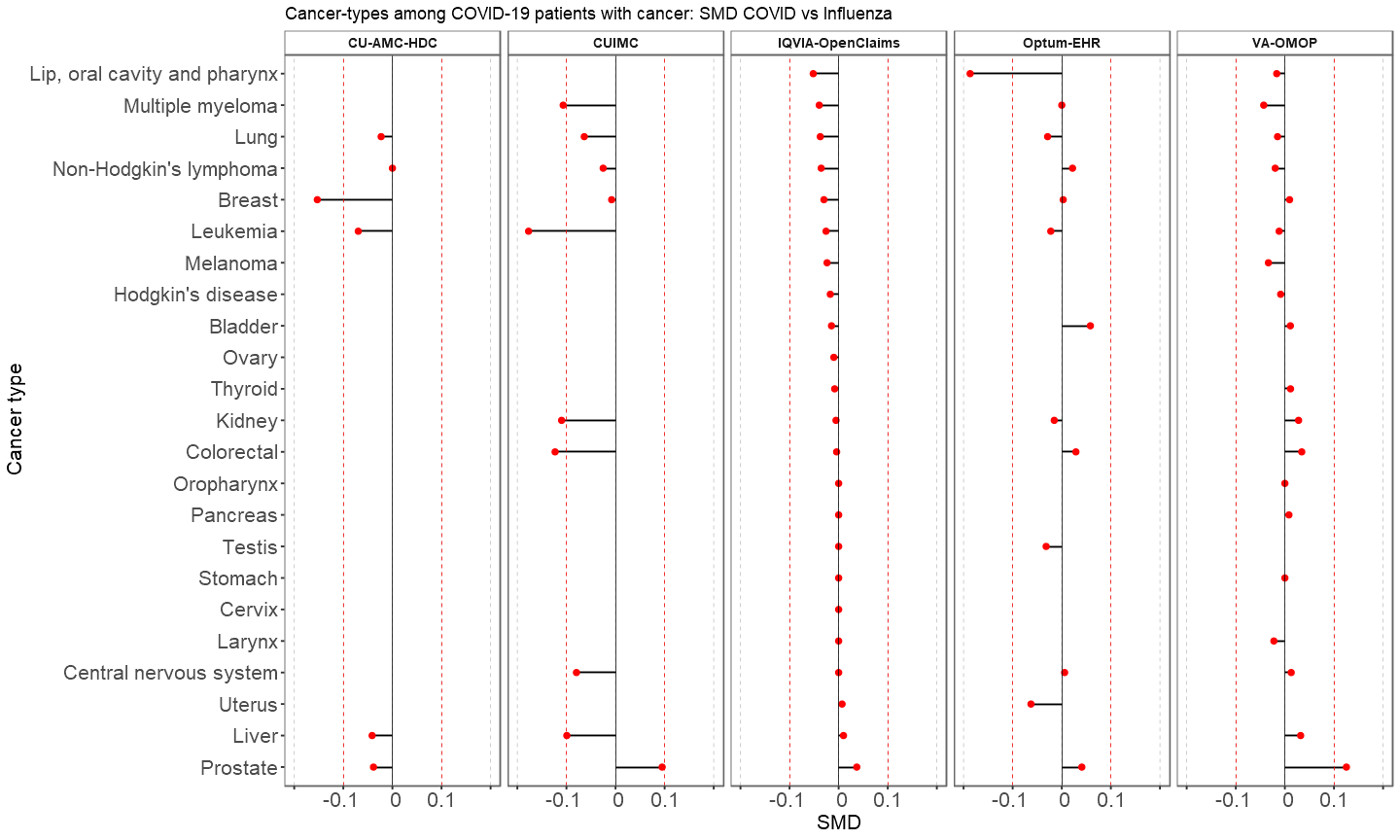

Notes: SMD calculated for cancer types meeting the minimum count required (5 individuals) in each database and cohort. Cancer types are ordered according to SMD descending values in IQVIA-OpenClaims. Abbreviations: COVID-19: Coronavirus Disease 2019; CU-AMC-HDC: Colorado University Anschutz Medical Campus Health Data Compass; CUIMC: Columbia University Irving Medical Center; VA-OMOP:Department of Veterans Affairs; SMD: Standardized mean difference

#### Appendix Table A8. Demographics, comorbidities and outcomes among patients with a history of cancer hospitalized with seasonal influenza 2017-2018

| **Characteristics, in % (95%CI)** | **CU-AMC-HDC**  **n= 219** | **CUIMC**  **n= 179** | **IQVIA-OpenClaims**  **n= 57,529** | **Optum-EHR**  **n= 473** | **VA-OMOP**  **n= 3,677** |
| --- | --- | --- | --- | --- | --- |
| Male | 53.0  (46.4-59.6) | 47.5  (40.2-54.8) | 47.7  (47.3-48.1) | 40.2  (35.8-44.6) | 95.8  (95.2-96.4) |
| **Antineoplastic agents** |  |  |  |  |  |
| Year prior | 40.2  (33.7-46.7) | 39.1  (31.9-46.3) | 24.0  (23.7-24.3) | 18.8  (15.3-22.3) | 23.7  (22.3-25.1) |
| Month prior | 32.0  (25.8-38.2) | 31.8  (25.0-38.6) | 13.9  (13.6-14.2) | 10.8  (8.0-13.6) | 14.4  (13.3-15.5) |
| **Comorbidities** |  |  |  |  |  |
| Asthma | 16.0  (11.1-20.9) | 34.1  (27.1-41.1) | 17.5  (17.2-17.8) | 28.3  (24.2-32.4) | 9.7  (8.7-10.7) |
| COPD | 46.6  (40.0-53.2) | 30.2  (23.5-36.9) | 40.2  (39.8-40.6) | 34.7  (30.4-39.0) | 64.6  (63.1-66.1) |
| Type 2 diabetes | 32.0  (25.8-38.2) | 57.5  (50.2-64.8) | 49.2  (48.8-49.6) | 39.5  (35.1-43.9) | 56.3  (54.7-57.9) |
| Hyperlipidemia | 42.9  (36.3-49.5) | 32.4  (25.5-39.3) | 44.6  (44.2-45.0) | 53.1  (48.6-57.6) | 60.5  (58.9-62.1) |
| Obesity | 47.9  (41.3-54.5) | 55.3  (48.0-62.6) | 27.5  (27.1-27.9) | 58.8  (54.4-63.2) | 48.7  (47.1-50.3) |
| Heart disease | 71.2  (65.2-77.2) | 79.3  (73.4-85.2) | 79.2  (78.9-79.5) | 67.2  (63.0-71.4) | 82.6  (81.4-83.8) |
| Hypertension | 74.9  (69.1-80.7) | 79.9  (74.0-85.8) | 87.4  (87.1-87.7) | 71.9  (67.8-76.0) | 92.6  (91.8-93.4) |
| Anxiety | 22.4  (16.9-27.9) | 11.7  (7.0-16.4) | 13.6  (13.3-13.9) | 25.6  (21.7-29.5) | 26.5  (25.1-27.9) |
| Dementia | 10.0  (6.0-14.0) | 16.2  (10.8-21.6) | 14.6  (14.3-14.9) | 5.5  (3.4-7.6) | 16.2  (15.0-17.4) |
| Depression | 6.4  (3.2-9.6) | 16.8  (11.3-22.3) | 12.0  (11.7-12.3) | 11.2  (8.4-14.0) | 17.2  (16.0-18.4) |
| Anemia | 39.3  (32.8-45.8) | 41.3  (34.1-48.5) | 32.6  (32.2-33.0) | 28.3  (24.2-32.4) | 34.4  (32.9-35.9) |
| Autoimmune condition | 14.6  (9.9-19.3) | 42.5  (35.2-49.8) | 35.1  (34.7-35.5) | 24.9  (21.0-28.8) | 34.7  (33.2-36.2) |
| Chronic kidney disease | 37.4  (31.0-43.8) | 42.5  (35.2-49.8) | 39.2  (38.8-39.6) | 33.8  (29.5-38.1) | 42.2  (40.6-43.8) |
| Chronic liver disease | 5.5  (2.5-8.5) | 4.5  (1.5-7.5) | 2.5  (2.4-2.6) | 3.4  (1.8-5.0) | 7.8  (6.9-8.7) |
| **Outcomes** |  |  |  |  |  |
| Death | - | - | - | - | 6.3  (5.5-7.1) |
| Intensive services | - | - | 3.8  (3.6-4.0) | 9.7  (7.0-12.4) | 12.9  (11.8-14.0) |
| ARDS | - | 14.0  (8.9-19.1) | 18.1  (17.8-18.4) | 24.7  (20.8-28.6) | 29.9  (28.4-31.4) |
| Cardiovascular events | - | 16.8  (11.3-22.3) | 13.3  (13.0-13.6) | 13.1  (10.1-16.1) | 22.2  (20.9-23.5) |
| Deep vein thrombosis | - | 6.1  (2.6-9.6) | 1.8  (1.7-1.9) | 2.3  (0.9-3.7) | 2.2  (1.7-2.7) |
| Pulmonary embolism | - | - | 1.4  (1.3-1.5) | 1.9  (0.7-3.1) | 2.0  (1.5-2.5) |
| AKI | - | 14.5  (9.3-19.7) | 7.6  (7.4-7.8) | 9.9  (7.2-12.6) | 11.8  (10.8-12.8) |
| Sepsis | - | 19.0  (13.2-24.8) | 16.8  (16.5-17.1) | 20.1  (16.5-23.7) | 21.9  (20.6-23.2) |

Note: - means data not available or below the minimum cell count required (5 individuals)

Abbreviations: CI: Confidence Interval; COPD: Chronic obstructive pulmonary disease; ARDS: Acute Respiratory Distress Syndrome; AKI: Acute Kidney Injury; SIDIAP: Information System for Research in Primary Care; CU-AMC-HDC: Colorado University Anschutz Medical Campus Health Data Compass; CUIMC: Columbia University Irving Medical Center; STARR-OMOP: Stanford Medicine Research Data Repository; VA-OMOP:Department of Veterans Affairs; SMD: Standardized mean difference

**Appendix Figure A5. Age distribution among patients with a history of cancer hospitalized with COVID-19 and with seasonal influenza**

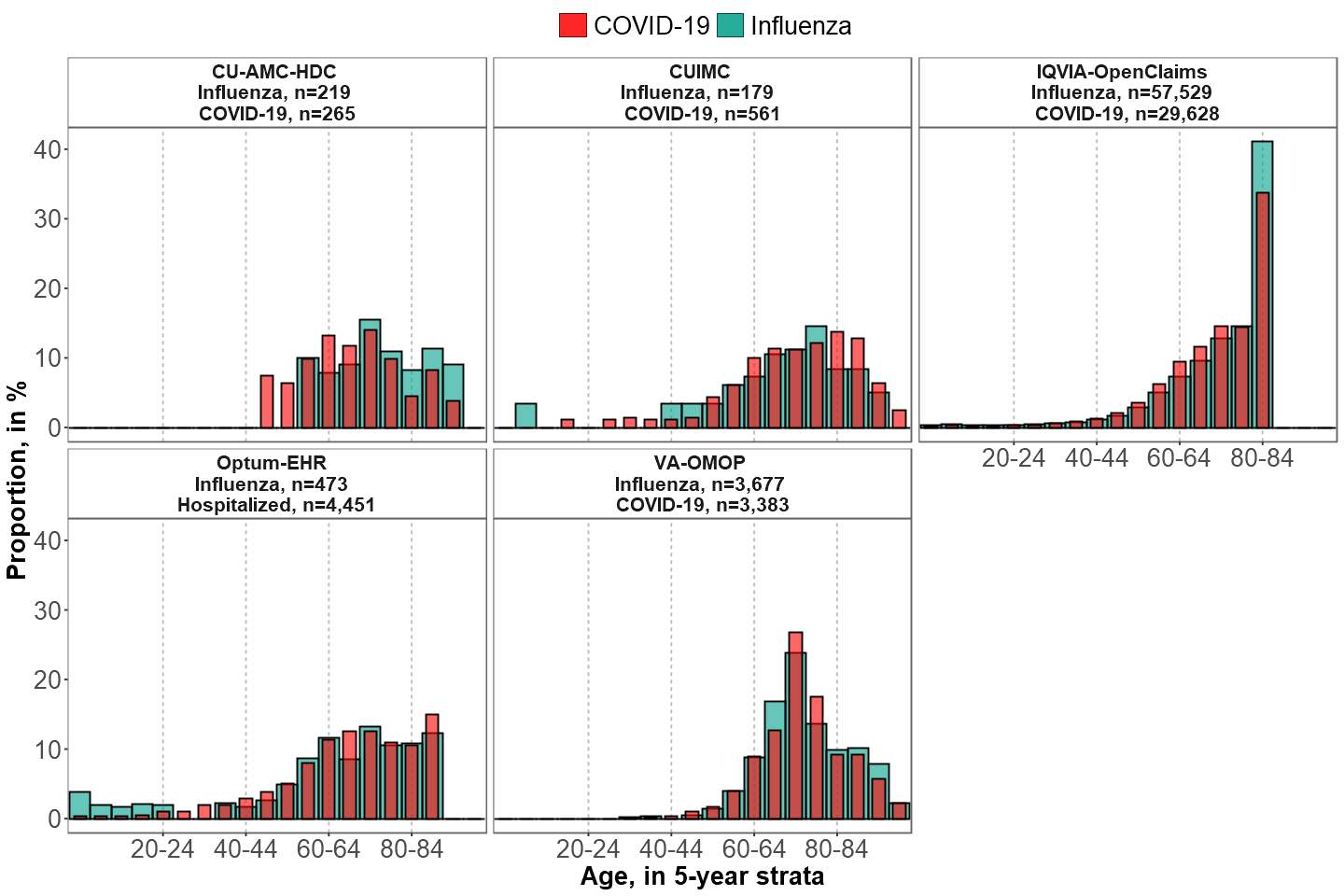

Note: In IQVIA-OpenClaims the age category 80-84 equals to “80 years or older”

Abbreviations: COVID-19: Coronavirus Disease 2019; SIDIAP: Information System for Research in Primary Care; CU-AMC-HDC: Colorado University Anschutz Medical Campus Health Data Compass; CUIMC: Columbia University Irving Medical Center; STARR-OMOP: Stanford Medicine Research Data Repository; VA-OMOP:Department of Veterans Affairs.
